# Privilege Is Not Enough: Evaluating Generalized Distillation for Normative Brain Age Modeling in Depression, Anxiety and Alcohol Use Disorder

**DOI:** 10.64898/2026.11.13.26368025

**Authors:** Johannes Nitsche, Julius Wiegert, Sajad Rezaei, Sebastian Volkmer, Sebastián Marty-Lombardi, Tobias Gradinger, Fabian Streit, Emanuel Schwarz

## Abstract

Models trained on rich multimodal data may depend on modalities unavailable at deployment. In generalized distillation, a teacher trained on such privileged data guides a student using only deployable features. We ask whether students inherit clinically meaningful properties beyond predictions in brain-age modeling, where brain-age gaps index deviations from normative aging and brain health. In simulations, distillation recovered a gap-severity association which an age-trained baseline missed. In UK Biobank analyses of depression, anxiety, and alcohol use disorder, students using structural MRI, functional MRI or biosocial data produced gaps closer to their teacher’s than matched baselines in 10 of 12 comparisons. Where teachers most exceeded baselines (anxiety in biosocial, alcohol use in functional MRI students), gap-severity associations rose toward their teacher’s. Our findings characterize conditions for successful distillation, i.e. clinical signal the teacher holds, the baseline misses and deployable features contain, and demonstrate their relevance in a real-life psychiatry application.

---

Despite extensive efforts to characterize the biology of mental illness, few markers have reached the individual-level predictive accuracy needed for clinical application. A central reason is that individual biomarkers typically show small and heterogeneous effects that do not map cleanly onto current diagnostic categories [1, 2]. Psychiatric research has therefore increasingly turned to machine learning (ML), which can integrate weak but distributed effects across many variables [3]. Interest has also grown in multimodal ML, because single modalities rarely capture the biological and clinical complexity of psychiatric disorders [4–7].

However, multimodal ML also creates substantial translational problems. First, multimodal data are fragmented across cohorts and often differ in acquisition protocols, preprocessing, and availability [8, 9]. Second, richer data collection increases cost [10], complexity, and participant burden [11], which limits routine use in clinical settings. Third, models that rely on a large and changing set of inputs are difficult to stabilize and validate for real-world deployment [12]. These issues weigh heavily in psychiatry, where reproducibility, external validity and clinical interpretability remain central challenges [13].

One way to reconcile these constraints is to exploit rich information during model development without requiring it at application. This idea is formalized in the paradigm of learning using privileged information (LUPI), where additional information is available only at training time [14–16]. By analogy to human instruction, the model with access to privileged information is called the teacher, and the deployable model it supervises the student [15]. The student must operate without privileged information at test time, ideally outperforming an identical model trained without a teacher. A structurally similar mechanism underlies knowledge distillation (KD) in deep learning, where the softened output distribution of a large or ensemble model serves as a training target for a smaller one [17]. Generalized distillation (GD) unifies both perspectives, transferring information across models and data representations [18]. Despite their theoretical appeal, these methods have seen little systematic evaluation in psychiatry, and it remains unclear whether privileged information yields practical benefit and, if so, under which conditions [19–21].

In the present study, we investigate GD through normative brain age modeling. In this approach, a model is trained to predict chronological age from biological data in a reference population; the difference between predicted and chronological age – the brain age gap (BAG) – quantifies individual deviation from normative aging [22, 23]. BAG has been widely investigated as a transdiagnostic index of brain health and is elevated, indicating an older-appearing brain, across a range of psychiatric and neurological conditions [24–26]. Normative brain age modeling offers two advantages for our purpose. First, it yields a single, interpretable, individual-level score whose clinical associations can be compared directly across models. Second, the same target can be predicted from different modalities, allowing teacher and student to learn from different data. The student does not need to use imaging. It can estimate an age gap from inexpensive, routinely collected cognitive, lifestyle, physical and sociode-mographic (biosocial) measures. Such non-imaging age gaps have themselves been associated with psychiatric and mortality outcomes [27, 28]. A biosocial student is therefore a meaningful recipient of a brain-age signal, rather than merely a cheaper approximation of the teacher. Brain age provides a well-characterized setting in which to evaluate GD, but the approach applies to other regression and classification problems in psychiatry.

This work pursues three aims. First, in simulated data (Fig. 1**a**), we construct a symptom-related component of brain aging that is uncorrelated with age and strongly present in the privileged features but weakly in the deployable ones, so that an age-trained model using only deployable features (the baseline) cannot recover it. This mimics psychiatric signal, which is typically weak and distributed across many features. We ask whether GD (Fig. 1**b**) can transfer this component to a student that, like the baseline, receives only deployable features at test time (Fig. 1**c**). Second, using neuroimaging (structural magnetic resonance imaging (sMRI) and resting-state functional MRI (rs-fMRI)) and biosocial data (Fig. 1**d**), we test whether GD makes the student’s raw (age-uncorrected) BAGs more teacher-like, i.e. more strongly correlated with the teacher’s BAGs across individuals, than the baseline model’s, moving beyond the binary-classification setting in which GD is typically studied. Third, we assess whether GD transfers the teacher’s associations between BAG and symptom severity in prevalent and burdensome mental and substance use disorders, and between BAG and cognition (Fig. 1**f**). With these aims, we ask not whether privileged information helps on average, but when it transfers and what is retained, linking the theory of GD to its practice in computational psychiatry.

**Fig. 1:**
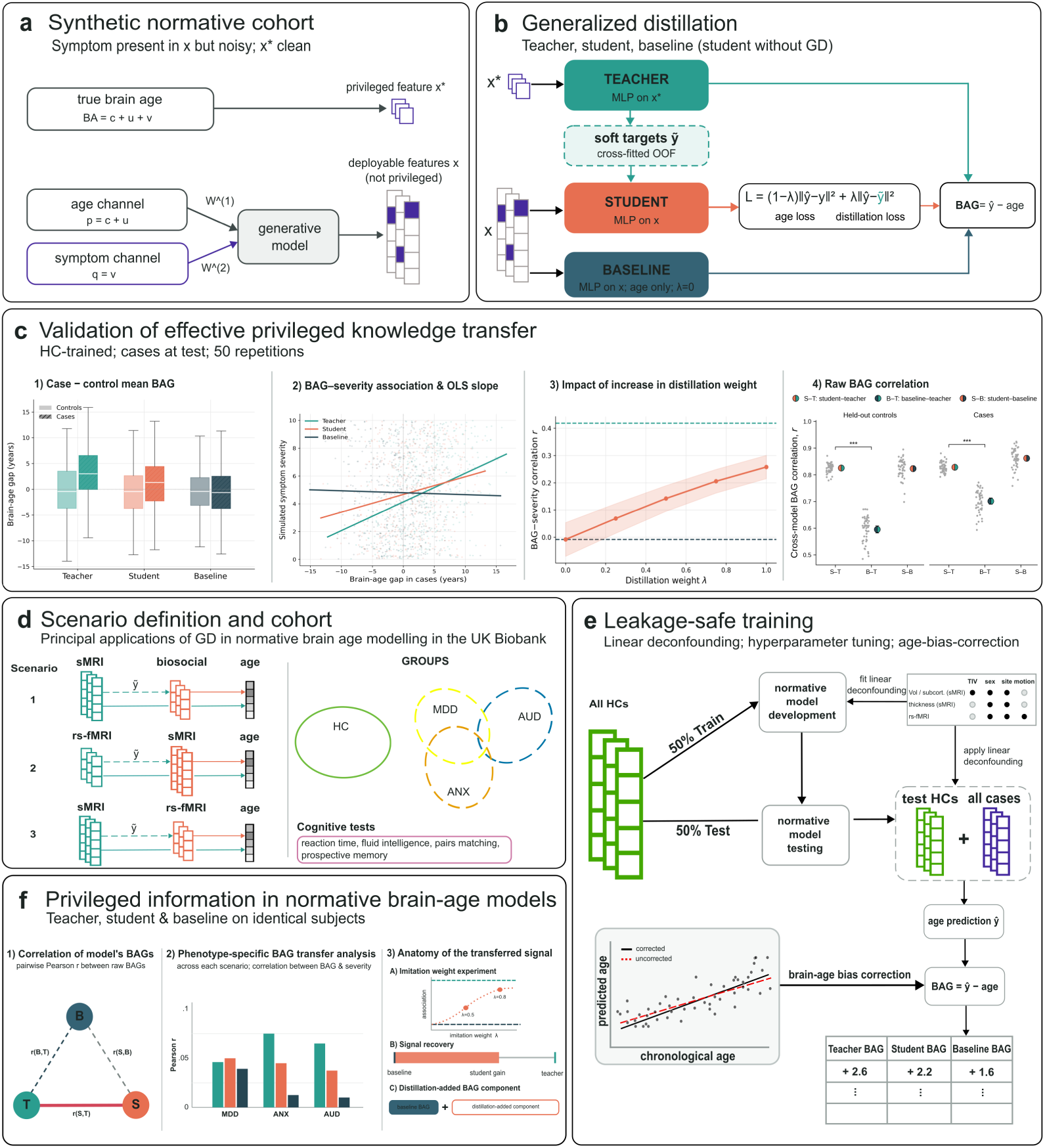
Schematic overview of the analysis pipeline for evaluating privileged-information transfer in normative brain-age modeling. **a,** We construct a synthetic cohort in which the true brain age is encoded in a privileged feature *x^∗^*, while the deployable features *x* carry the same age- and symptom-related signal but in a noisily entangled, non-privileged form. **b**, A teacher model trained on the privileged feature *x^∗^* emits cross-fitted soft targets *ỹ*. A student model trained on the deployable features *x* then learns from a mixture of the age and a distillation loss, whereas a baseline model sees only the age target (*λ* = 0). **c**, Models are trained on controls only and evaluated on held-out controls and on cases across 50 repetitions. For student and baseline, we quantify the case-control BAG difference and the BAG-severity association, and examine how both change as the distillation weight *λ* is increased. **d**, We define three application scenarios in the UKB dataset according to which modality plays the privileged (teacher) versus the deployable (student and baseline) role, and examine them across the groups (HC, MDD, ANX, AUD) and cognitive measures. **e**, HCs are split 50/50; deconfounding, hyperparameter tuning and age-bias correction are fit on the training HCs only before BAGs are computed for the held-out HCs and for all cases. **f**, We dissect the transfer in the real data by comparing teacher, student and baseline BAGs on identical subjects, through their pairwise correlations, phenotype-specific BAG-severity associations, and a decomposition of the distillation-added signal. Abbreviations: GD, generalized distillation; BAG, brain age gap; HC, healthy control; AUDIT-C, Alcohol Use Disorders Identification Test-Consumption; AUD, alcohol use disorder; PHQ-9, Patient Health Questionnaire-9; MDD, major depressive disorder; GAD-7, Generalized Anxiety Disorder-7; GAD, generalized anxiety disorder; ANX, anxiety disorder; ICD-10, International Classification of Diseases, 10th revision.

## Results

We evaluated GD first in simulation and then in the UK Biobank (UKB, [29]) (Fig. 1). The teacher used privileged information throughout, the student received it only through the teacher’s training outputs, and the baseline matched the student except for the distillation term, so their difference isolates the effect of distillation. All models shared a leakage-safe normative pipeline fit on training controls (Fig. 1**e**; Methods). After confirming the mechanism in simulation, we evaluated three teacher-student scenarios in the UKB, each representing a common challenge in clinical deployment or cross-modal translation. We asked whether distilled students reproduce their teacher’s BAGs and inherit the teacher’s associations between the gap and symptom severity or cognition. The clinical groups comprised individuals with major depressive disorder (MDD), anxiety disorder (ANX) and alcohol use disorder (AUD), defined by the International Classification of Diseases, Tenth Revision (ICD-10); within each group, symptom severity was measured with the Patient Health Questionnaire-9 (PHQ-9), the Generalized Anxiety Disorder-7 (GAD-7) and the Alcohol Use Disorders Identification Test-Consumption (AUDIT-C), respectively (Table 1; Supplementary Section S1.3.5).

**Table 1:** Demographics for the two UKB samples. The Structural-functional sample includes participants with complete sMRI and rs-fMRI data. In the Structural-biosocial sample, participants with complete sMRI and with cognitive (fluid intelligence, reaction time, pairs matching and prospective memory) and sociodemographic (e.g. smoking status, sleep, education) data are included. Clinical questionnaire-based rows describe severity strata for reference; association analyses used the ICD-10 groups. Columns report group size (N), mean age (±s.d.) and percentage female. Abbreviations: HC, healthy controls; AUDIT-C, Alcohol Use Disorders Identification Test-Consumption; AUD, alcohol use disorder; PHQ-9, Patient Health Questionnaire-9; MDD, major depressive disorder; GAD-7, Generalized Anxiety Disorder-7; GAD, generalized anxiety disorder; ANX, anxiety disorder; ICD-10, International Classification of Diseases, 10th revision. Subgroups are not mutually exclusive

| **Group** | **Criterion** | **N** | **Age** *μ*(±*σ*) | **Females (%)** |
|---|---|---|---|---|
| *Structural-functional sample* |  |  |  |  |
| HC |  | 12,705 | 65 (±7) | 53 |
| AUDIT-C (8) | AUDIT-C sum = 8 | 1,071 | 62 (±7) | 36 |
| AUDIT-C (9) | AUDIT-C sum = 9 | 792 | 62 (±7) | 29 |
| AUDIT-C (≥ 10) | AUDIT-C sum ≥ 10 | 847 | 62 (±7) | 27 |
| AUD (ICD-10) | ICD-10 | 935 | 65 (±7) | 43 |
| PHQ-9 (mild) | PHQ-9 sum ∈ [5, 9] | 3,224 | 62 (±8) | 63 |
| PHQ-9 (moderate) | PHQ-9 sum ∈ [10, 14] | 735 | 61 (±7) | 62 |
| PHQ-9 (moderately severe) | PHQ-9 sum ∈ [15, 19] | 257 | 60 (±7) | 66 |
| PHQ-9 (severe) | PHQ-9 sum ≥ 20 | 128 | 57 (±6) | 70 |
| MDD (ICD-10) | ICD-10 | 715 | 62 (±8) | 64 |
| GAD-7 (mild) | GAD-7 sum ∈ [5, 9] | 2,817 | 62 (±8) | 66 |
| GAD-7 (moderate) | GAD-7 sum ∈ [10, 14] | 533 | 62 (±8) | 65 |
| GAD-7 (severe) | GAD-7 sum ∈ [15, 21] | 269 | 59 (±7) | 68 |
| ANX (ICD-10) | ICD-10 | 649 | 63 (±8) | 66 |
| *Structural-biosocial sample* |  |  |  |  |
| HC |  | 7,132 | 64 (±7) | 52 |
| AUDIT-C (8) | AUDIT-C sum = 8 | 954 | 62 (±7) | 35 |
| AUDIT-C (9) | AUDIT-C sum = 9 | 714 | 62 (±7) | 30 |
| AUDIT-C (≥ 10) | AUDIT-C sum ≥ 10 | 777 | 62 (±7) | 27 |
| AUD (ICD-10) | ICD-10 | 595 | 64 (±8) | 40 |
| PHQ-9 (mild) | PHQ-9 sum ∈ [5, 9] | 1,936 | 62 (±8) | 62 |
| PHQ-9 (moderate) | PHQ-9 sum ∈ [10, 14] | 421 | 60 (±7) | 63 |
| PHQ-9 (moderately severe) | PHQ-9 sum ∈ [15, 19] | 153 | 59 (±7) | 65 |
| PHQ-9 (severe) | PHQ-9 sum ≥ 20 | 62 | 56 (±6) | 74 |
| MDD (ICD-10) | ICD-10 | 400 | 61 (±8) | 63 |
| GAD-7 (mild) | GAD-7 sum ∈ [5, 9] | 1,677 | 62 (±8) | 65 |
| GAD-7 (moderate) | GAD-7 sum ∈ [10, 14] | 313 | 61 (±8) | 62 |
| GAD-7 (severe) | GAD-7 sum ∈ [15, 21] | 141 | 59 (±7) | 73 |
| ANX (ICD-10) | ICD-10 | 371 | 62 (±7) | 64 |

### Simulation-based privileged brain-age signal analysis

By construction, the symptom-linked component was uncorrelated with age and only weakly present in the deployable features, so the age-prediction objective alone could not recover it from them. The privileged teacher’s BAG was elevated in cases relative to controls (mean case–control difference ΔBAG*_T_* = 4.31 *±* 0.36 yr, mean *±* SD over 50 repetitions; Fig. 2**a**) and was associated with symptom severity (*r_T_* = 0.42 *±* 0.04, *p_F DR_* = 5.72 *×* 10*^−^*^46^; Fig. 2**b**). The deployable baseline was effectively blind to this signal, meaning its BAG neither differed between cases and controls (ΔBAG*_B_* = *−*0.27 *±* 0.45 yr) nor tracked severity (*r_B_* = 0.01*±*0.06, *p_F DR_* = 0.36). Distilled on the teacher’s soft age predictions, the student tested on the deployable features recovered a substantial fraction of both. The student’s BAG differed between cases and controls (ΔBAG*_S_* = 2.08 *±* 0.35 yr) and was associated with severity (*r_S_*= 0.22 *±* 0.04, *p_F DR_* = 9.52 *×* 10*^−^*^36^), roughly half of the teacher’s association.

**Fig. 2:**
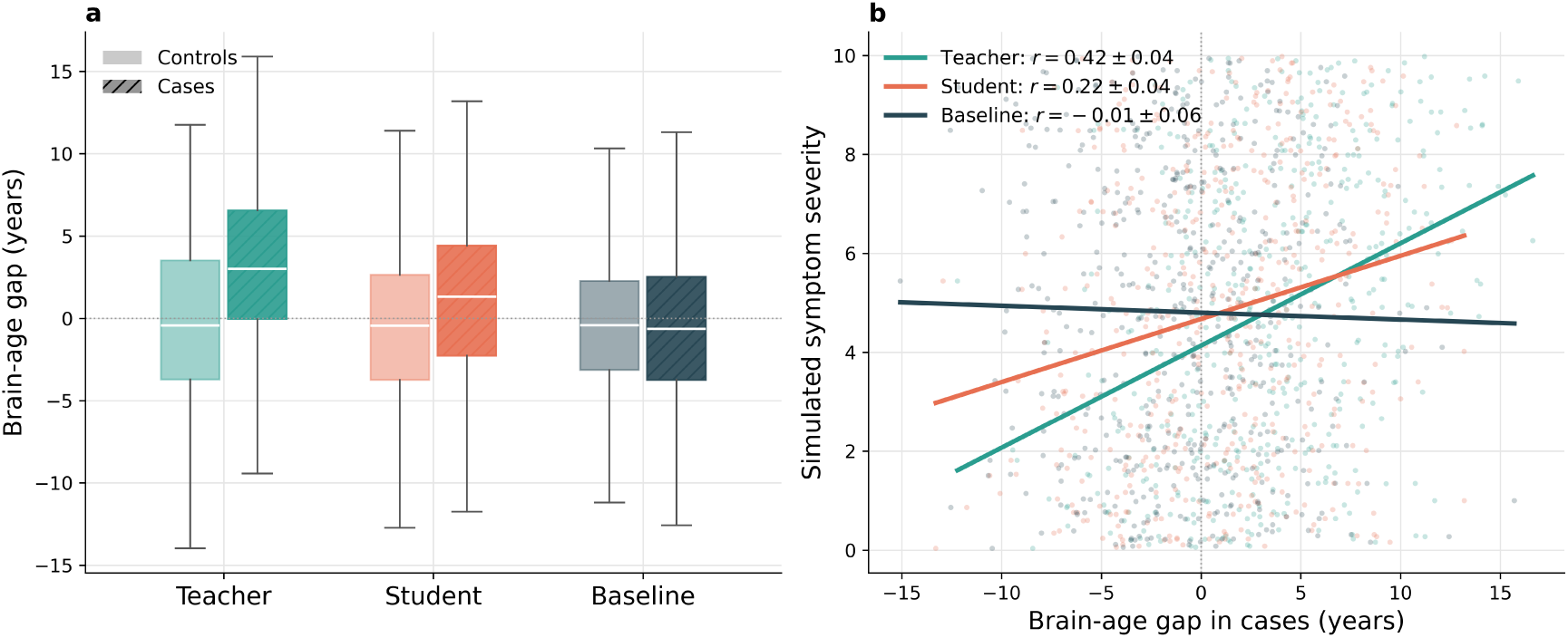
Generalized distillation transfers a privileged, symptom-linked brain-age gap to a deployable student in simulation. **a,** Brain age gap (BAG) for held-out controls and cases under the teacher (privileged feature *x^∗^*), the baseline and the distilled student (both tested with deployable features *x*). The case elevation in BAG present for the teacher is absent for the baseline and partially restored for the student. **b,** Symptom severity as a function of BAG in cases, with the BAG as the predictor; per-model regression lines and Pearson correlations *r* (mean ± standard deviation over 50 repetitions) are shown. Boxplots and scatter depict a representative split.

Across the 50 repetitions, all pre-specified effects were significant. The student was reliably more teacher-like than the baseline, its BAGs correlating more strongly with the teacher’s than the baseline’s did (*r_ST_ − r_BT_* = 0.13, 95% CI [0.12, 0.14], *d_z_* = 3.1, *p*_FDR_ *<* 0.001) and the student’s BAG-severity correlation exceeded the baseline’s (mean difference = 0.22, 95% CI [0.21, 0.24], *d_z_* = 4.9, *p*_FDR_ *<* 0.001; Supplementary Table S10). The transfer scaled with the distillation weight, i.e. increasing *λ* from 0 (which recovers the baseline) to 1 moved the student monotonically toward the teacher on both the BAG-severity correlation and the case-control separation (Supplementary Fig. S3), confirming the distillation term of the loss as the source of privileged information transfer.

### Evaluation of generalized distillation on UK Biobank data

We applied GD in three teacher-student scenarios (Fig. 1**d**; Methods). An imaging teacher distilled into a biosocial student (scenario 1), and two scenarios that use the same two imaging modalities but reverse which one is privileged (scenarios 2 and 3).

#### Correlation analysis between models’ BAGs

We compared the age-uncorrected BAGs of the teacher, student and baseline in healthy controls (HCs) and in the three ICD-10 case groups (MDD, ANX and AUD). For each group we plotted the three pairwise correlations as a triangle whose vertices are the models, placing more strongly correlated models closer together (Fig. 3, balanced KD weighting; construction in Supplementary Section S1.7).

**Fig. 3:**
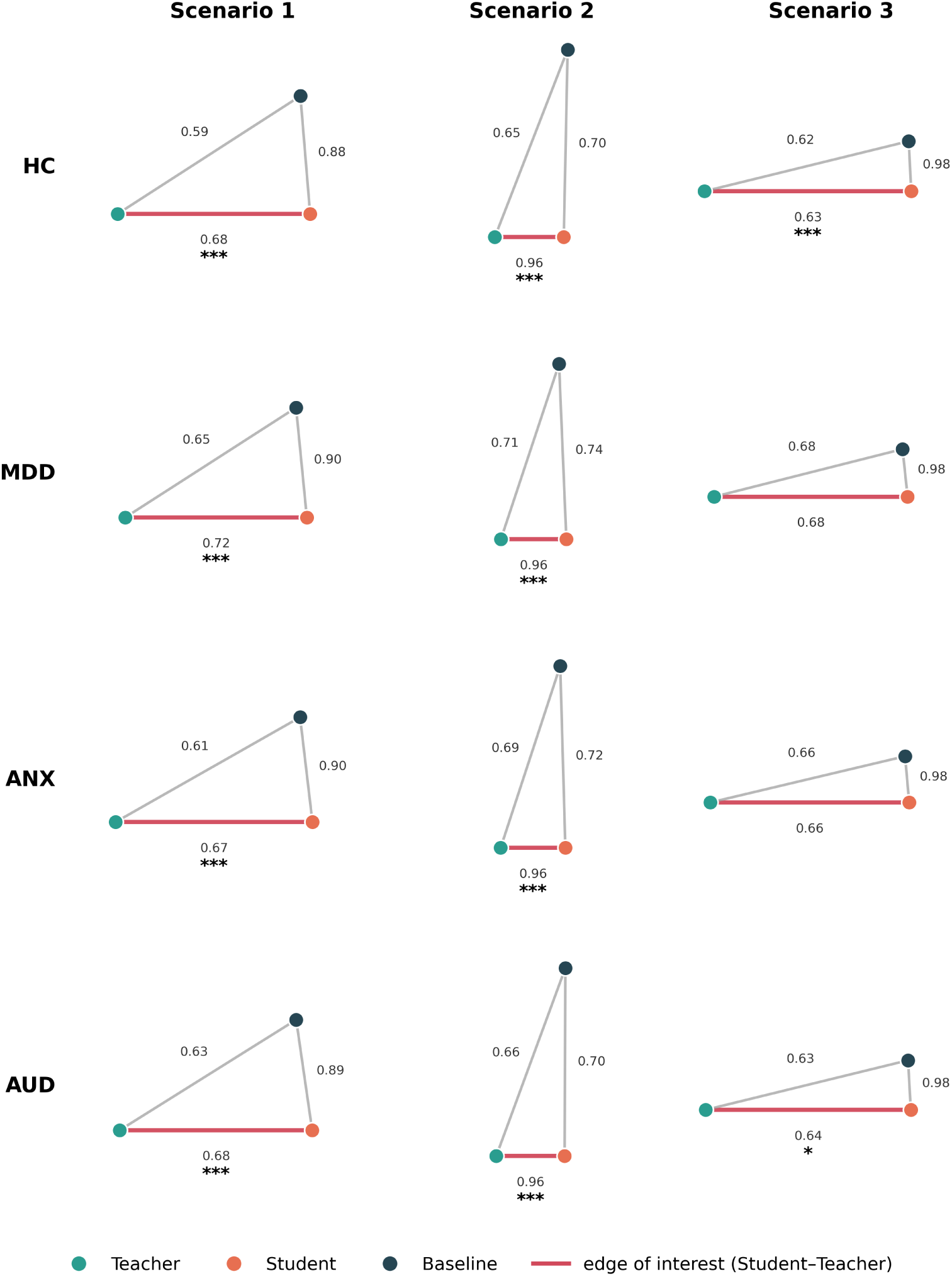
Cross-model comparison of the raw BAGs, shown as a distance geometry. Each panel places the teacher, student and baseline models as the vertices of a triangle for one group (rows: HCs and the ICD-10 MDD, ANX and AUD diagnostic groups) and one distillation scenario (columns). Vertices that sit closer have more strongly correlated BAGs; each edge is labeled with its Pearson *r*. The student-teacher edge (red) is the teacher-likeness contrast of interest. Asterisks beneath its coefficient denote that the student’s BAGs are significantly more teacher-like than the baseline’s (i.e. *r*_ST_ *> r*_BT_) by a one-sided paired bootstrap with Benjamini-Hochberg correction across the twelve group × scenario cells (*^∗^ p_F DR_ <* 0.05, *^∗∗∗^ p_F DR_ <* 0.001). Construction details of the figure can be found in Supplementary Section S1.7. Abbreviations: BAG, brain age gap; HC, healthy control; AUD, alcohol use disorder; MDD, major depressive disorder; ANX, anxiety disorder; ICD-10, International Classification of Diseases, 10th revision.

##### Scenario 1 (teacher sMRI *→* student biosocial)

The student and baseline produced closely correlated BAGs (*r_SB_ ≈* 0.88–0.90), which showed lower correlations with the sMRI teacher’s (*r_ST_≈* 0.67–0.72, *r_BT_ ≈* 0.59–0.65). The student was nonetheless more teacher-like than the baseline in every group (*r_ST_ > r_BT_*, *p_F DR_ <* 0.001), the expected signature of transfer.

##### Scenario 2 (teacher rs-fMRI *→* student sMRI)

The student-teacher BAG correlation was by far the strongest (*r_ST_ ≈* 0.96 in all rows), exceeding both the student-baseline (*r_SB_ ≈* 0.70–0.74) and baseline-teacher (*r_BT_ ≈* 0.65–0.71) correlations (*p_F DR_ <* 0.001), even though the student and baseline share the sMRI modality. Distillation pulled the student’s BAG almost onto the rs-fMRI teacher’s.

##### Scenario 3 (teacher sMRI *→* student rs-fMRI)

With the modalities reversed, the student’s and baseline’s BAGs were almost identical (*r_SB_≈* 0.98) and both correlated only weakly with the teacher’s (*r_ST_, r_BT_ ≈* 0.62–0.68). The student was more teacher-like than the baseline in HCs (*p_F DR_ <* 0.001) and in the AUD group (*p_F DR_ <* 0.05), but not in the MDD or ANX group.

The two imaging scenarios were strongly asymmetric. At balanced weighting the rs-fMRI teacher reshaped the sMRI student almost completely (scenario 2), whereas the sMRI teacher barely moved the rs-fMRI student from its baseline (scenario 3). The same correlation-analysis on simulated data gave a similar pattern to scenario 1 (Supplementary Fig. S2). In scenarios 1 and 3 the student still resembled its baseline more than the teacher, yet remained more teacher-like than the baseline in six of eight instances. We next asked whether this shared signal carries clinically relevant structure.

#### Recovery of teacher-defined BAG–symptom associations

We investigated whether the student recovered the teacher’s phenotype-specific signal (its within-group BAG-severity association), scoring severity with PHQ-9 in MDD, GAD-7 in ANX and AUDIT-C in AUD. Because scenario 2’s teacher showed no elevated BAG–symptom association (Supplementary Fig. S5), its student had no phenotype-specific signal to recover. Fig. 4 therefore shows only scenarios 1 and 3.

**Fig. 4:**
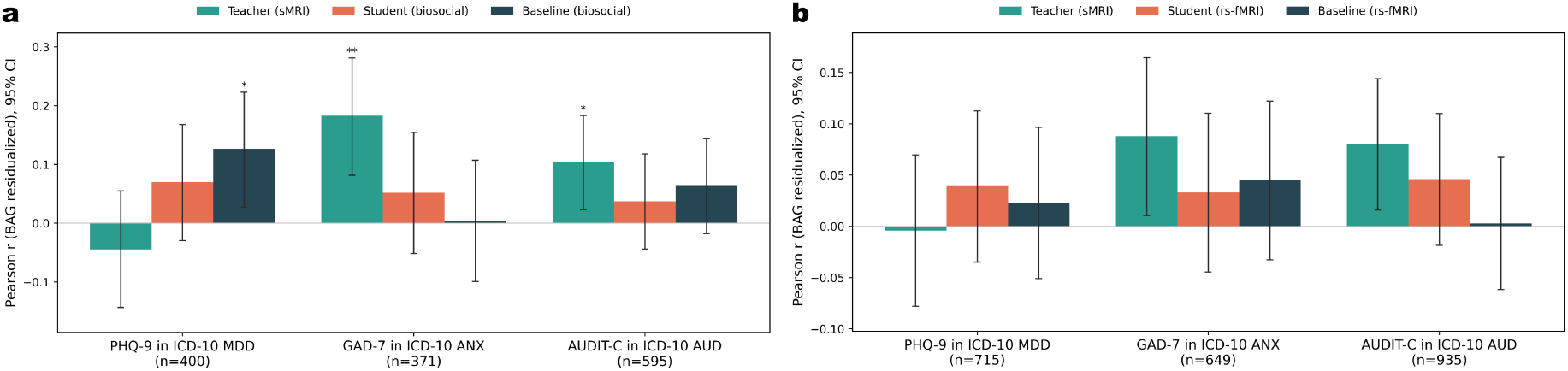
Phenotype-specific transfer of the teacher’s BAG-symptom-severity association. Semi-partial Pearson correlations (95% CI) between each model’s BAG and symptom severity, within the matched MDD, ANX and AUD groups (*λ* = 0.5). **a,** Scenario 1: The sMRI teacher’s anxiety association is partly recovered by the biosocial student. **b,** Scenario 3: The sMRI teacher shows an elevated BAG–AUDIT-C association, which is partially transferred to the rs-fMRI student relative to its baseline; in this scenario, no association survives FDR correction. The panels use different y-axis ranges, so bar heights are not directly comparable across scenarios. BAGs were adjusted for confounders. CIs were obtained by Fisher *z*-transformation; asterisks mark associations significantly different from zero after Benjamini–Hochberg correction across models and subgroups within each scenario: *^∗^ p_F DR_ <* 0.05, *^∗∗^ p_F DR_ <* 0.01.

##### Scenario 1 (teacher sMRI *→* student biosocial)

The sMRI teacher carried the only significant symptom associations in the ANX (*r_T_ ≈* 0.18, *p_F DR_ <* 0.01) and AUD (*r_T_ ≈* 0.10, *p_F DR_ <* 0.05) subgroups, whereas the biosocial baseline carried a significant association for depression severity (PHQ-9 in MDD, *r_B_ ≈* 0.14, *p_F DR_ <* 0.05; Fig. 4**a**). GD partially transferred the teacher’s anxiety signal, raising the student’s BAG-symptom severity association (*r_S_ ≈* 0.05) above its baseline’s (*r_B_ ≈* 0.004), though not to significance. No comparable transfer was found for MDD or for the AUD association, where the student’s associations roughly halved compared to the baseline. A biosocial student can thus partly recover the sMRI teacher’s symptom signal, but only for specific phenotypes.

##### Scenario 2 (teacher rs-fMRI → student sMRI)

The fMRI teacher’s associations between BAG and PHQ-9, GAD-7 and AUDIT-C were weak and non-significant (Supplementary Fig. S5). The student’s teacher-like BAGs therefore yielded no clinical or cognitive gain.

##### Scenario 3 (teacher sMRI → student rs-fMRI)

Here the privileged modality carried transferable structure for the BAG–AUDIT-C association instead of GAD-7 as in scenario 1. In the AUD group, where the sMRI teacher showed a strong association and the rs-fMRI baseline essentially none, the student rose above baseline toward the teacher (Fig. 4**b**; *r_T_ ≈* 0.08, *r_S_ ≈* 0.05, *r_B_ ≈* 0.003). None of the associations survived FDR correction, and the elevation was confined to AUD. The moderate shift of the student’s raw BAG (*r_ST_ ≈* 0.64) together with its corrected BAG–AUDIT-C association suggests that GD transferred a small, targeted part of the teacher’s sMRI signal rather than the bulk of its estimate.

#### Anatomy of the transferred signal

We characterized how the signal transferred, adding cognitive outcomes, through three complementary views (Fig. 5). The transfer grew with imitation strength (**a**), it was bounded by how much signal the teacher actually held over baseline (**b**), and it was carried by the component of the student’s BAG that GD added on top of the baseline (**c**).

**Fig. 5:**
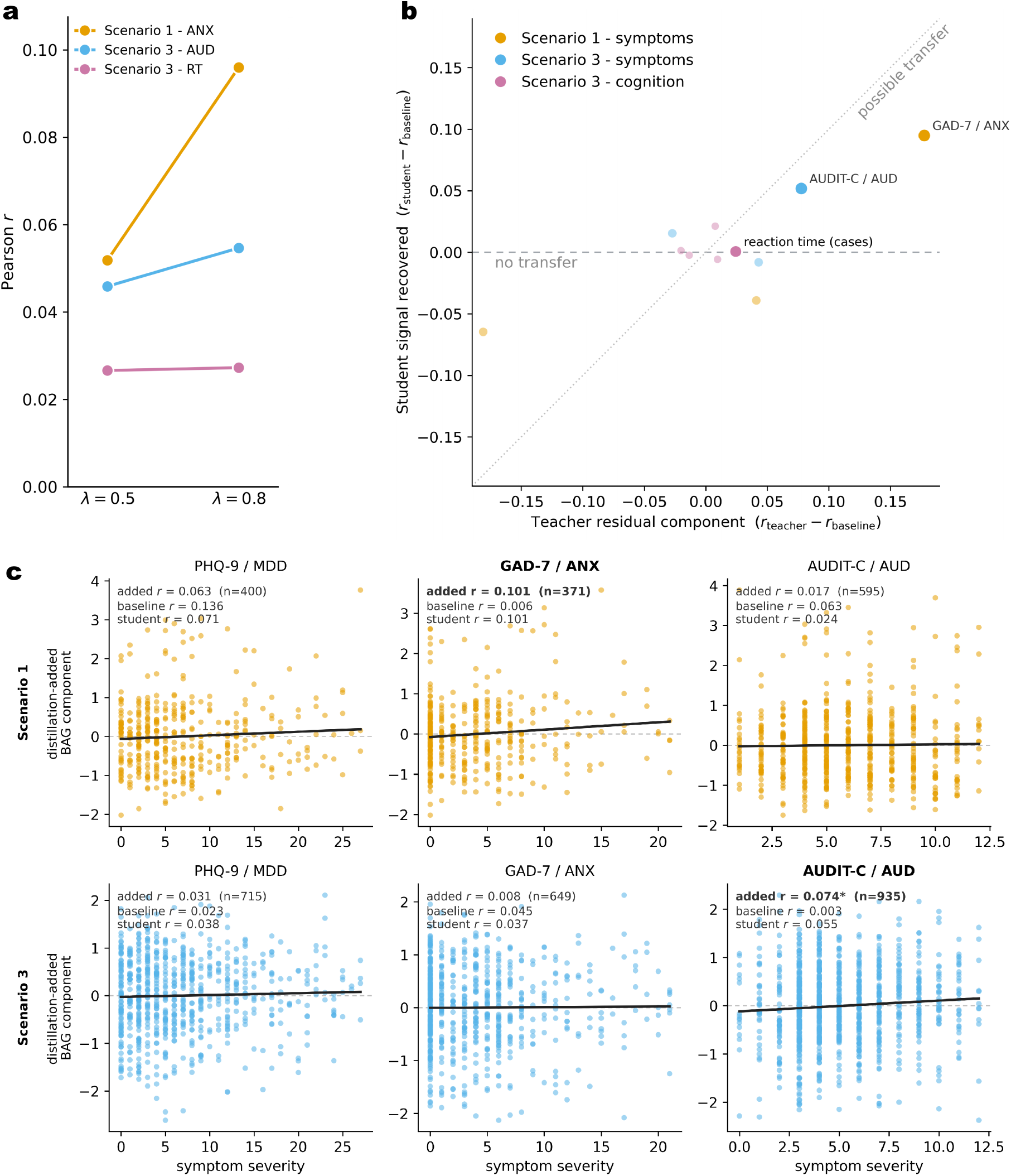
Anatomy of the transferred signal. **a,** Semi-partial Pearson *r* between the student’s BAG and symptom severity (and reaction time) at *λ* = 0.5 and *λ* = 0.8. For the two transferring symptom associations, the correlation was larger under stronger imitation; reaction time (RT, in cases) was included as a non-symptom control and showed no meaningful increase. **b,** Student signal recovered (*r_S_− r_B_*) against the residual teacher signal (*r_T_ − r_B_*) for symptom and cognitive outcomes. The dashed line marks no transfer and the dotted diagonal marks full recovery of the teacher signal; recovery tracks the available teacher signal, which is elevated for the AUD and ANX symptom associations but small for RT. **c,** The distillation-added BAG component (student BAG adjusted for confounders and the baseline BAG) plotted against symptom severity for scenario 1 (top) and scenario 3 (bottom). “Added *r*” is the correlation of this component with the respective questionnaire score; *^∗^ p*_FDR_ *<* 0.05, Benjamini– Hochberg corrected across the two transferring cells. Panels **b** and **c** show the *λ* = 0.8 run. All correlations are semi-partial: BAGs were adjusted for confounders, whereas outcomes were not adjusted. Abbreviations: BAG, brain age gap; AUDIT-C, Alcohol Use Disorders Identification Test-Consumption; AUD, alcohol use disorder; PHQ-9, Patient Health Questionnaire-9; MDD, major depressive disorder; GAD-7, Generalized Anxiety Disorder-7; ANX, anxiety disorder.

##### Stronger teacher imitation increases the transferred signal

For the two scenarios in which the student produced teacher-like associations between predicted BAG and symptom severity (scenario 1 - ANX, *n* = 371; scenario 3 - AUD, *n* = 935), we compared the strength of that association at the balanced imitation weight (*λ*_1_ = 0.5) and at a higher weight (*λ*_2_ = 0.8). As the simulation predicted (Supplementary Fig. S3), the semi-partial correlation between the student’s confounder adjusted BAG and symptom severity increased in both scenarios (Fig. 5**a**). In scenario 1 it rose from *r_λ_*_1_ = 0.053 to *r_λ_*_2_ = 0.097, and a one-sided Steiger test suggested a significant increase after BH FDR correction (*Z* = 2.27, *p* = 0.011, *p_F DR_* = 0.034; Supplementary Table S13). In scenario 3 the increase was smaller, from *r_λ_*_1_ = 0.049 to *r_λ_*_2_ = 0.058, and did not reach significance (*Z* = 0.82, *p* = 0.21, *p_F DR_* = 0.31). The same *λ*-sweep applied to a non-symptom outcome (reaction time in cases, *n* = 7552) produced only a marginal change (*r_λ_*_1_ = 0.028 to *r_λ_*_2_ = 0.029; *Z* = 0.16, *p* = 0.43).

##### Transfer is bounded by the available teacher signal

Across symptom and cognitive outcomes, the amount of signal the student recovered over its baseline scaled with how much signal the teacher held over that same baseline (Fig. 5**b**). Where the teacher carried a substantial privileged association – anxiety in scenario 1 (Δ*_T B_* = *r_T_ − r_B_* = 0.18) and alcohol use in scenario 3 (Δ*_T B_* = 0.08) – the student recovered a meaningful fraction of it (ANX: student gain *≈* 0.10, *≈* 54% of the teacher’s advantage; AUD: student gain *≈* 0.05, *≈* 67%). By contrast, for three cognitive measures the teacher’s advantage over baseline was small (reaction time Δ*_T B_* = 0.02; pairs matching and fluid intelligence Δ*_T B_ ≤* 0.01), so there was little signal to transfer and the student recovered essentially none of it (reaction time: student gain *≈* 0.001). Recovery fractions have wide bootstrap intervals that include zero (Supplementary Table S14) and are indicative only, but the pattern is consistent.

##### The severity signal localizes to the distillation-added BAG component

Finally, we asked whether the transferred association lives in the part of the student’s BAG that distillation actually adds, rather than in variance the student would have carried anyway. We residualized the student’s BAG against the confounders and the baseline’s BAG, and correlated the resulting distillation-added component with symptom severity (Fig. 5**c**). In the two phenotype-scenario cells where transfer was observed, the added component itself carried a positive severity association: scenario 1 ANX (added *r* = 0.101, *p_F DR_* = 0.056, *n* = 371) and scenario 3 AUD (added *r* = 0.074, *p_F DR_* = 0.048, *n* = 935). The corresponding non-transferring cells were near zero or dominated by the baseline (e.g. scenario 1 AUD added *r* = 0.017; scenario 3 ANX added *r* = 0.008), and where the baseline already carried the association (MDD in scenario 1 (baseline *r* = 0.136)) the transferred component added little. A severity signal in the distillation-added component suggests additional evidence of a distillation effect. We note that only the scenario 3 AUD association reached significance; the numerically larger scenario 1 ANX association did not, and all effect sizes are modest.

## Discussion

Distillation from a privileged teacher consistently made deployable students resemble it, yet beyond simulated data, this resemblance carried over the teacher’s clinical associations only under two identifiable conditions. First, the teacher had to encode phenotype-specific structure that a matched baseline missed (teacher advantage); second, the deployable features had to be able to express that structure (expressibility). Where both conditions were satisfied, the student showed a trend toward recovering part of the teacher’s advantage, an effect that grew with imitation weight and resided in the distillation-added BAG component. Where either was unmet, distillation altered the student’s predictions without adding clinical signal. In our experiments, teacher-likeness was therefore an insufficient proxy for transfer. The value of privileged information has to be assessed on the downstream property it is intended to carry.

The simulation established that the mechanism operates when teacher advantage and expressibility hold by construction. The age-trained baseline ignored a symptom-linked component that was uncorrelated with age and weakly present in the deployable features, yet the distillation term alone conveyed roughly half of the teacher’s case-control separation and BAG-severity association to the student. This transfer grew monotonically with the imitation weight and vanished at *λ* = 0, isolating the GD loss as its source [15, 18]. In UK Biobank data the same signature appeared for anxiety with a biosocial student (scenario 1) and for alcohol use with an rs-fMRI student (scenario 3). In both, the student’s association moved from a baseline near zero toward the teacher’s, rose with imitation weight (significantly only for anxiety) and was carried by the distillation-added BAG component rather than by variance shared with the baseline. Across outcomes, the student’s gain over the baseline scaled with the teacher’s. These conditions are consistent with LUPI theory, in which privileged information improves a student only insofar as it carries target-relevant information that the student does not learn from its own deployable data [15].

Scenario 2 showed the opposite case, imitation without transfer. Distillation moved the sMRI student’s raw BAG almost onto the rs-fMRI teacher’s (*r_ST_* = 0.96, against *r_BT_* between 0.65 and 0.71), but the teacher carried no symptom or cognition association, so the student gained none. Imitation was not free in any scenario. Every student’s mean absolute error (MAE) was 0.2 to 1.7 years higher than its baseline’s (Supplementary Table S11). Less accurate age models have been shown to be more disease-sensitive [30]. A loss in age-prediction accuracy may therefore be acceptable where it is accompanied by a gain in clinical signal. In scenarios 1 and 3, this cost came with a nominal gain for one phenotype each; in scenario 2, it yielded no clinical benefit. Two properties plausibly explain the asymmetry between the imaging scenarios. The rs-fMRI teacher was a weak age predictor, and the raw BAG of a weak predictor is dominated by chronological age through regression toward the mean [31, 32]. Part of the scenario 2 agreement likely reflects this shared age dependence rather than transferred deviation structure, since the student also lost age accuracy. In addition, the sMRI student had more input features and capacity than the rs-fMRI student (Supplementary Table S1) and could more readily reproduce a foreign BAG geometry. Teacher-likeness thus reflects teacher prediction accuracy and student capacity as much as transfer. This parallels evidence that a student’s fidelity to its teacher and its own performance can diverge [21].

The form of the distillation objective also mattered. Soft-label distillation, which matches the teacher’s predicted age distribution for each subject, produced no improvement over the baseline on UK Biobank data, likely because these distributions are dominated by the age signal that the supervised loss term already supplies. In the simulation, by contrast, the teacher’s predictions deviated from chronological age mainly through the symptom-linked component, so matching each subject’s prediction transferred that component directly. Relational knowledge distillation [33] instead matches normalized distances between the teacher’s BAGs and is therefore unaffected by shifting or rescaling them, a property it shares with the correlations used to measure BAG–severity associations. This may explain why it conveyed the severity signal on UK Biobank data. Where clinical signal resides in deviations from a normative prediction, distillation objectives should target that deviation structure rather than the prediction itself.

Two practical implications follow. First, a candidate teacher should be screened for case-relevant signal beyond a matched baseline before a LUPI pipeline is built around it. Second, our analyses form a reusable audit for distillation studies. The matched baseline isolates what distillation changed, the teacher’s advantage over it bounds what can be recovered, and the decomposition into baseline-shared and distillation-added components localizes the transferred signal. The two conditions also guide the choice of method where GD alone does not suffice. Where expressibility is lacking, the remedy lies in the choice of deployable modality rather than in the training procedure, since no training-time method, including modality synthesis [34], can supply structure absent from the deployable features. Where the teacher holds no advantage over the baseline, as in scenario 2, reliability-aware distillation, which down-weights unreliable teacher outputs in favor of the ground-truth target, is more appropriate [19]. Signal distributed across several modalities may be better exploited by multimodal teachers with auxiliary alignment terms [35–37], shared representations from contrastive, geometric or latent-factor objectives [38, 39], or test-time adaptation with privileged information [40] than by a single privileged channel. GD is the simplest member of this family, requiring only a modified loss and, given its conditional payoff, we recommend evaluating it alongside these alternatives.

This work has several limitations. The UK Biobank is older, healthier and less symptomatic than clinical populations, which likely attenuates the BAG–symptom associations, and severity relied on brief self-report questionnaires. Effects were modest: two remained significant after FDR correction, namely the distillation-weight-dependent strengthening of the BAG–GAD-7 association and the added-component association with AUDIT-C, and recovery fractions had wide confidence intervals. The null result for depression is best explained by missing teacher advantage, as the sMRI teacher held no depression association beyond the baseline. Normative models trained only on controls may be unable to encode structure that emerges solely in cases, leaving it unavailable for transfer. Expressibility was inferred from the transfer pattern rather than measured independently. Finally, transfer was assessed only through the BAG, and supervised phenotype prediction may behave differently. Natural extensions include teachers from modalities with strong clinical loadings such as PET imaging [41], case-inclusive or severity-aware normative models, criteria for choosing the imitation weight by clinical sensitivity in independent data, and replication across clinical cohorts.

In conclusion, privileged information during training is not enough. GD can carry a privileged brain-age signal into models that rely only on cheap, accessible inputs, cleanly in simulation and partially in real psychiatric data. Its value depends on whether the deployable features can capture teacher knowledge that age-based supervision alone cannot reveal, and agreement with the teacher does not by itself demonstrate that the student acquired this knowledge. Evaluating GD on the downstream property it is meant to transfer, rather than on imitation, will show where it improves on a model trained without privileged information.

## Methods

### Simulation study

#### Synthetic normative cohort

We simulated a normative cohort from a fully specified generative model in which a symptom-linked component of brain aging, uncorrelated with age, is strongly expressed in a privileged feature *x^∗^* but only weakly in the deployable features *x*.

Each sample has a chronological age, drawn uniformly over 20 to 80 years. We define true brain age as chronological age plus two deviations. The first is a generic deviation, present in every sample and unrelated to symptoms. The second is a symptom-linked deviation. In controls the symptom-linked deviation is centered on zero and unrelated to symptom severity. In the case group we sample a bounded latent scalar representing symptom severity, independently of chronological age. The symptom-linked deviation increases with the severity score, resulting in a higher latent BAG in more severe cases. Severity is the external criterion against which the estimated BAGs are evaluated.

The simulation generates two feature sets. The deployable set has eight features. Each is a noisy linear mixture of two latent channels with fixed loadings. One channel is the age channel, equal to chronological age plus the generic deviation. The other is the symptom channel, equal to the symptom-linked deviation. The age channel dominates the variance of every deployable feature, and the symptom channel accounts for only a small share of it. The symptom channel is also uncorrelated with chronological age. A model trained to predict chronological age from the deployable features therefore has no reason to recover it. The privileged set contains a single feature, true brain age itself, which carries the symptom-linked deviation directly, without the attenuation and measurement noise that obscure it in the deployable features. This deviation is therefore the privileged information. It is available to the teacher through the privileged feature but is nearly absent from the student’s features. The full generative equations and parameter values are given in Supplementary Methods S1.1.

Training followed the normative convention. All models were trained on controls only. Cases were introduced only at test time. Each repetition comprised newly simulated, non-overlapping cohorts of 300 training controls, 500 held-out controls and 500 cases. We ran 50 repetitions. No hyperparameters were tuned; all models used a single fixed configuration, identical across conditions and repetitions (Supplementary Section S1.1.4).

#### Normative models and generalized distillation

Three regressors predicting chronological age were compared. The teacher was a multilayer perceptron trained on the privileged feature *x^∗^*; as the privileged model it retained access to *x^∗^* at test time and is therefore not deployable. In practice, *x^∗^* stands for a measurement that informs model development but is unavailable at the point of care – for example, molecular imaging (amyloid or tau PET, which captures disease-related neuropathology [41]), where routine clinical assessment provides only sMRI or questionnaires. The baseline was a multilayer perceptron trained on the deployable features *x*, such as sMRI, against chronological age alone. The student shared the baseline architecture and deployable inputs but was trained by GD of the teacher. The teacher’s soft targets were its standardized out- of-fold (OOF) predictions on the training controls (5-fold cross-fitting), which approximate recovered brain age and thus encode the per-control offset that the chronological-age label omits. Cross-fitting ensures that each control’s soft target is produced by a teacher that did not see that control, so no in-sample teacher fit leaks into the student. The student minimized the regression form of the distillation objective,

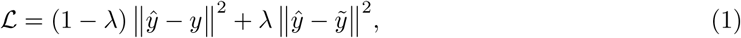

where *y*^ is the student prediction, *y* the standardized chronological age and *ỹ* the teacher soft target, with imitation weight *λ*. Features and the age target were standardized using training-control statistics. All simulation results are reported at *λ* = 0.8; the effect of *λ* was examined by sweeping it from 0 to 1 (Supplementary Fig. S3).

Symptom severity was generated independently of chronological age, so the BAG-severity association was not confounded by the simulated age dependence of the gap; the standard linear age-bias correction was therefore not applied here but remains advisable on empirical data [23].

#### Outcome measures and statistical analysis

For each repetition, we calculated the Pearson correlation between BAG and simulated illness severity among cases. We report the ordinary-least-squares (OLS) slope from regressing illness severity on BAG among cases – following the brain-age convention, with BAG as the predictor and illness severity as the dependent variable – and the mean case-control difference in BAG (ΔBAG) across held-out controls and cases. Estimates are reported as the mean ± s.d. over the 50 repetitions. Three prespecified hypotheses were tested with one-sided paired *t*-tests across repetitions, i.e. that the student’s per-participant BAGs agree with the teacher’s more closely than the baseline’s do (test subject-level correlation with teacher BAG); that the student’s BAG is increased in cases relative to controls; and that the student’s BAG-severity correlation exceeds the baseline’s. *P* -values were corrected across the three tests by the Benjamini-Hochberg (BH) procedure [42] (adjusted values are denoted *p_F DR_*); effect sizes are Cohen’s *d_z_* for paired samples with 95% *t*-based confidence intervals. Full simulation parameter settings, model hyperparameters and the mechanistic rationale are given in Supplementary Methods S1.1.

### UK Biobank study

#### Cohort and group definitions

The UKB is a population-based cohort of approximately 500,000 participants aged 40-69 years at recruitment [29], with sMRI data available for 40,325 individuals and rs-fMRI within- and between-network connectivity measures for 40,336 individuals. Individuals with neurodegenerative diseases (see Supplementary Table S3 for ICD-10 codes) were excluded. We further excluded participants with missing clinical-symptom questionnaire data (PHQ-9 [43], GAD-7 [44], and AUDIT-C [45]). We then defined two analysis samples, each requiring complete confounder information. The Structural-functional sample comprised participants with complete sMRI and rs-fMRI data. Here, confounders were age, sex, scan site, total intracranial volume (TIV), and mean head motion during rs-fMRI scans. Every participant in this sample was required to have both imaging modalities because the cross-modal analyses use sMRI and rs-fMRI reciprocally, each serving as teacher in one direction and as student in the other. The Structural-biosocial sample comprised participants with complete sMRI together with cognitive (fluid intelligence, reaction time, pairs matching and prospective memory) and sociodemographic (e.g. smoking status, sleep, education) data, and excluded rs-fMRI. Its confounders were age, sex, scan site and TIV. Group distributions and demographics for both samples are summarized in Table 1. Current depressive, anxiety, and alcohol-use symptoms were assessed using the PHQ-9, GAD-7, and AUDITC, respectively, and participants were stratified into severity strata using cut-offs (PHQ-9: mild 5–9, moderate 10–14, moderately severe 15–19, severe ≥20 [43]; GAD-7: mild 5–9, moderate 10–14, severe 15–21 [44]; AUDIT-C: 8, 9, and ≥ 10 [45]). Healthy controls (HCs) were defined as PHQ-9 *<* 5, GAD-7 *<* 5, AUDIT-C *<* 8, with no self-reported or ICD-10-diagnosed MDD or ANX, and no ICD-10 diagnosis of AUD, schizophrenia, bipolar disorder, attention-deficit hyperactivity disorder, borderline personality disorder, or Alzheimer’s disease (for ICD-10 codes, see Supplementary Table S4). In addition to questionnaire-based definitions, ICD-10 code-based diagnoses for MDD, ANX, and AUD were used to define diagnostic groups [46].

#### Neuroimaging data processing

Structural (T1-weighted) and rs-fMRI data were acquired and preprocessed by UK Biobank following its standardized imaging protocol and processing pipeline [47, 48].

Structural features were FreeSurfer-derived morphometric measures from the UK Biobank brain imaging pipeline [48, 49]. These comprised 27 subcortical, ventricular and infratentorial volumes plus five corpus-callosum subdivisions from the automated subcortical segmentation, and cortical thickness and volume for the 34 Desikan-Killiany regions of each hemisphere [50], for 168 structural features per participant.

Resting-state functional connectivity was derived from Schaefer 100-parcel node time series, each parcel assigned to one of the seven Yeo intrinsic connectivity networks [51, 52]. After discarding the first five non-steady-state volumes, a parcel-level Pearson correlation matrix was computed for each participant and averaged within the network structure to summarize connectivity at the network level, yielding seven within-network values (mean correlation among parcels of the same network) and 21 across-network values (mean correlation between parcels of each network pair), for 28 functional connectivity features per participant.

Acquisition parameters, quality-control, imaging-derived phenotypes, and the corresponding UK Biobank field identifiers are reported in Supplementary Methods S1.3.

#### Choice and processing of biosocial features

The biosocial feature set was assembled from routinely collected questionnaire responses and simple physical measurements, with no imaging, laboratory assays or specialist assessment required. These data could be, therefore, in principle, obtained at low cost and minimal participant burden. This feature set comprises cognitive measures (fluid intelligence, reaction time, pairs matching and prospective memory), lifestyle and health variables (smoking status, sleep duration, insomnia, body mass index and doctor-diagnosed diabetes), physical measurements (waist-to-hip ratio and dominant-hand grip strength), and sociodemographic indicators (educational attainment, household income, Townsend deprivation index [53] and number in household).

Candidate variables were chosen for their established correlation with chronological age and brain-related measures. Fluid intelligence and processing or reaction speed decline across adulthood [54], and grip strength, adiposity, sleep and cardiometabolic measures change systematically with age. These variables have documented associations with brain structure and related outcomes, including cognitive performance and regional gray-matter volume [55], grip strength [56], body mass index (BMI) and waist-to-hip ratio [57], sleep duration and insomnia [58], and socioeconomic indicators such as education and income [59]. Biomedical, lifestyle and cognitive factors of this kind have also been associated with neuroimaging-derived brain age [60]. Finally, variables were recorded for a substantial fraction of participants, ensuring adequate overlap with the imaging sub-sample.

The full list of UK Biobank fields (Supplementary Table S8) and the derivation, recoding, and cleaning steps for each variable are given in Supplementary Methods S1.5.

#### Teacher–student scenarios

We evaluated GD in three scenarios, each defined by the privileged modality available to the teacher during training and the deployable modality shared by student and baseline. In scenario 1 (Structural-biosocial sample), an sMRI teacher supervised a student trained on the biosocial features, whose prediction residual is thus a biosocial age gap. In scenario 2 (Structural-functional sample), an rs-fMRI teacher supervised an sMRI student. Scenario 3 inverted this pairing (sMRI teacher, rs-fMRI student) and served as a directional control rather than a deployment setting.

Each scenario comprised a teacher, a student and a matched baseline, fit on identical subjects with identical standardization. The baseline matched the student in architecture, hyperparameters, features, age target, and training and evaluation subjects, and differed only in omitting the distillation term from its loss. Distillation used the relational knowledge distillation objective (RKD [33]; Supplementary Section S1.2.2); soft-label distillation is described in Supplementary Section S1.2.1.

### Exploratory association analysis

#### Cross-model correlation of the raw brain age gaps

To test whether distillation shifted the student’s predictions toward the teacher’s, we compared the raw (age-uncorrected) BAGs of teacher, student and baseline at the balanced weighting (*λ* = 0.5; Fig. 3). We define a model’s *teacher-likeness* as the Pearson correlation of its raw BAGs with the teacher’s across subjects. The raw BAGs of any two age models correlate even without distillation, partly through their shared age dependence [31]. The student’s teacher-likeness (*r_ST_*) was therefore compared with that of the matched baseline (*r_BT_*), which differs from the student only in omitting the distillation term. We further compared *r_ST_* with the student–baseline correlation (*r_SB_*) to test whether the student resembled the teacher more than the baseline with which it shares its deployable features.

For each group (held-out HCs, MDD, ANX, AUD) and scenario, the three pairwise correlations were computed on the shared complete-case frame, with Fisher-*z* 95% confidence intervals (See Supplementary Fig. S4). Both hypotheses (*r_ST_ > r_BT_* and *r_ST_ > r_SB_*) were tested with a one-sided paired bootstrap (5,000 resamples of subjects), in which each resample entered both correlations so that their dependence was preserved. Benjamini-Hochberg correction was applied separately to each hypothesis across the twelve group *×* scenario cells.

#### Brain age gap and its association with outcomes

Motivated by evidence that the BAG tracks symptom burden and cognition within psychiatric groups, at times in a symptom-domain-specific way [61–63], we tested each model’s bias-corrected BAG for association with the symptom severity and cognition. Symptom scales were tested within their matched ICD-10 group (PHQ-9 in MDD, GAD-7 in ANX, AUDIT-C in AUD), and cognitive measures (fluid intelligence, reaction time, prospective memory and pairs matching) in all cases^1^. Associations were quantified as semi-partial Pearson correlations: each model’s BAG was residualized by OLS on the confounders of the respective sample (see Cohort and group definitions; full design in Supplementary Section S1.6.2), whereas the outcome was left unadjusted. This downstream adjustment follows brain-age convention and is applied in addition to the fold-wise feature residualization used during model fitting. Teacher, student and baseline were evaluated on the same complete-case subjects, so that their associations are directly comparable. Confidence intervals and two-sided *p*-values were derived from the Fisher-*z* transform with *n* - *k* - 3 degrees of freedom for *k* confounders, and Benjamini-Hochberg correction was applied across models and groups within each scenario. We denote the BAG–outcome associations of teacher, baseline and student by *r_T_*, *r_B_* and *r_S_*.

#### Phenotype-specific transfer and its scaling with imitation weight

Transfer was defined per outcome, i.e. per symptom scale (PHQ-9, GAD-7, AUDIT-C) or cognitive measure, as an elevation of the student’s BAG–outcome association above its distillation-free baseline toward a teacher that itself carried a non-trivial association (operationalized in Supplementary Section S1.6.2 as the teacher’s advantage over baseline and the student’s recovery fraction). The teacher, baseline and student correlations are reported together for every scenario and outcome. The BAG– outcome associations of teacher, baseline and student (Fig. 4) are reported at the balanced weighting (*λ*_1_ = 0.5). The signal-recovery and decomposition analyses (Fig. 5**b**,**c**) used the transfer-heavy run (*λ*_2_ = 0.8), in which teacher imitation dominates the loss. To test whether transfer strengthens as the imitation weight increases, motivated by the monotone *λ*-response of the simulation (Supplementary Section S2.1.3), we compared, on the same subjects, the student’s association at the balanced and transfer-heavy (*λ*_2_ = 0.8) weightings (Fig. 5**a**). This test was restricted to the associations identified as transferring, namely anxiety in scenario 1 and alcohol use in scenario 3, with reaction time as a non-symptom control. Both correlations are computed on the same subjects with the same outcome, and the *λ*_1_- and *λ*_2_-student BAGs are themselves highly correlated (*r >* 0.93; Supplementary Table S13), so the two correlations are not independent. We therefore compared them with Steiger’s test for two dependent correlations that share one variable [64], one-sided for an increase (*r_λ_*_2_ *> r_λ_*_1_), and applied Benjamini-Hochberg correction across the tested associations (anxiety, alcohol use and reaction time).

#### Signal recovery and localization of the transferred component

Two analyses characterized how the signal transferred. Both used the *λ*_2_-student and the matched baseline of that run. To quantify how much of the teacher’s privileged signal the student recovered, each association was expressed relative to the matched baseline. We define the teacher advantage as Δ*_T B_* = *r_T_* − *r_B_*, the student gain as *r_S_* −*r_B_*, and the recovery fraction is their ratio, (*r_S_* −*r_B_*)*/*Δ*_T B_*. The recovery fraction was evaluated only where the teacher advantage was positive, since the ratio is unstable as the denominator approaches zero. Its 95% confidence interval was obtained from a percentile bootstrap over subjects (1000 resamples) in which teacher, baseline and student were resampled jointly and re-residualized on every draw (Supplementary Table S14).

To test whether the transferred association resides in what distillation adds rather than in variance the student would carry regardless, we isolated the distillation-added component of the student BAG as the residual of the confounder-residualized student BAG after regression on the confounder-residualized baseline BAG, computed on identical subjects. It’s semi-partial correlation with symptom severity (added *r*) was tested against zero with Fisher-*z* intervals. Benjamini-Hochberg correction was applied across the two phenotype–scenario cells in which transfer had been identified in the preceding association analysis (scenario 1 ANX, scenario 3 AUD); the remaining cells are reported descriptively. A severity association present in the added component but absent from the baseline indicates a distillation effect rather than variance shared with the baseline (Supplementary Section S1.8).

#### Model training, deconfounding and evaluation

The procedure below was applied identically across all three scenarios, each a pairing of a privileged teacher modality with a deployed student modality. As feature correction is keyed to modality rather than to teacher or student role, the same residualization rules hold whichever modality supplies privileged or deployable information. Each model predicts age by classification over a one-year age grid with Gaussian-smoothed targets, taking predicted age as the grid expectation. Within each training split of a cross validation (CV) fold, each feature was linearly residualized on its modality-specific confounds (where defined) and z-scored, with both the residualization coefficients and the z-scoring statistics estimated on the training samples alone, then applied unchanged to the corresponding validation, normative holdout and case samples. Structural volumes and subcortical measures were residualized on TIV, sex and site; cortical thickness on sex and site. The resting-state across-network connectivity features were residualized on mean absolute head motion, sex and site (see Supplementary Section S1.6). Where applicable, questionnaire and cognitive features were z-scored without residualization.

HCs were partitioned once into a development sample (50% of HCs) and a normative holdout (the other half) by stratified sampling on age and sex; the holdout was neither used for tuning nor seen during training and was scored exactly once. Within the development sample we ran ten-fold CV, with each training portion further split into train and validation subsets for early stopping. Teacher supervision of the student was itself leakage-controlled. The student’s distillation targets on any training row were the teacher’s inner-cross-validated out-of-fold (OOF) predictions, so that no subject ever supervised the student through a teacher that had been trained on it, and no information was shared across outer folds.

CV with early stopping was used only to fix a common epoch budget (the median best epoch across folds); models were then refit at that fixed budget in a cross-fitting pass over the full development sample to yield OOF predictions. The BAG bias correction (a linear regression of the gap on chronological age, see [32]) was estimated on these development OOF predictions alone and applied forward, unchanged, to the normative holdout and to cases. Predictive accuracy (MAE, RMSE, Pearson *r*) was computed on the raw predicted age. The age-bias correction was applied only to the gaps carried forward into the association analyses. It does not by itself adjust for the remaining confounders; those are handled separately at the association stage.

Hyperparameters were selected in a two-stage procedure that used a development-set split seed aligned to the training run. The teacher was tuned first and frozen; the student and matched baseline were then tuned over the same grid of learning rate, weight decay and dropout, selecting on the pooled development OOF MAE averaged over seeds. The distillation weight (*λ* ∈ [0,1]) was not tuned. It was fixed per run and was increased systematically to investigate its effect on knowledge transfer. The baseline inherited the student’s selected settings, so that student-versus-baseline isolates the effect of distillation rather than of hyperparameter differences. As hyperparameters were selected per run, each *λ* run has its own matched baseline.

#### Use of large language models

We used Claude Opus 4.8 to assist with drafting and editing portions of the manuscript and to generate initial Python code for data preprocessing and figure generation. All generated text was reviewed and edited by the authors, who take full responsibility for its content. All generated code was manually reviewed, tested, and revised by the authors before use.

## Ethics declaration

This study was approved by the Ethics Committee II of the University of Heidelberg Medical Faculty Mannheim (protocol no. 2025-827). UKB data collection were approved by the relevant local ethics committees in the United Kingdom, respectively, and were conducted in accordance with the Declaration of Helsinki. All UKB participants provided written informed consent.

## Data availability

Access to UKB data requires application through the registration and application portal (http://ukbiobank.ac.uk/register-apply).

## Acknowledgments

We thank the participants and scientists involved in making the UKB resource available (http://www.ukbiobank.ac.uk/). This study was conducted under UKB application number 162313.

## Author contributions

J.N. conceived and designed the study, performed all analyses, developed the code and wrote the original draft of the manuscript. S.M.-L. preprocessed the resting-state fMRI data. J.W. contributed to data curation. E.S. supervised the project and acquired funding. All authors contributed to reviewing and editing the manuscript and approved the final version.

## Code availability

All analyses were conducted in Python (version 3.12). All analysis scripts and the packages used are available via GitHub at https://github.com/nitschejohannes/privileged-brain-age.

## Footnotes

1 Association analyses between BAG and cognition were conducted only in scenarios 2 and 3, because cognitive testing was part of the biosocial student’s feature set in scenario 1.

## References

[1] Carvalho, A. F. et al. Evidence-based umbrella review of 162 peripheral biomarkers for major mental disorders. Translational Psychiatry 10, 152 (2020). URL https://www.nature.com/articles/s41398-020-0835-5.

[2] Schmaal, L. The curse and opportunity of heterogeneity in the pursuit of psychiatric biomarkers. World Psychiatry 22, 271–272 (2023). URL https://onlinelibrary.wiley.com/doi/abs/10.1002/wps.21085.

[3] Bzdok, D. & Meyer-Lindenberg, A. Machine Learning for Precision Psychiatry: Opportunities and Challenges. Biological Psychiatry. Cognitive Neuroscience and Neuroimaging 3, 223–230 (2018).

[4] Zhou, Z. et al. Machine learning methods in psychiatry: a brief introduction. General Psychiatry 33, e100171 (2020).

[5] Khoo, L. S., Lim, M. K., Chong, C. Y. & McNaney, R. Machine Learning for Multimodal Mental Health Detection: A Systematic Review of Passive Sensing Approaches. Sensors 24, 348 (2024).

[6] Koutsouleris, N. et al. Multimodal Machine Learning Workflows for Prediction of Psychosis in Patients With Clinical High-Risk Syndromes and Recent-Onset Depression. JAMA psychiatry 78, 195–209 (2021).

[7] Gordon, J. A., Dzirasa, K. & Petzschner, F. H. The neuroscience of mental illness: Building toward the future. Cell 187, 5858–5870 (2024).

[8] Acosta, J. N., Falcone, G. J., Rajpurkar, P. & Topol, E. J. Multimodal biomedical AI. Nature Medicine 28, 1773–1784 (2022). URL https://www.nature.com/articles/s41591-022-01981-2.

[9] Tognin, S. et al. Corrigendum to: Towards Precision Medicine in Psychosis: Benefits and Challenges of Multimodal Multicenter Studies-PSYSCAN: Translating Neuroimaging Findings From Research into Clinical Practice. Schizophrenia Bulletin 48, 722 (2020). URL https://pmc.ncbi.nlm.nih.gov/articles/PMC9077412/.

[10] Etkin, A., Powell, J. & Savitz, A. J. Opportunities for use of neuroimaging in de-risking drug development and improving clinical outcomes in psychiatry: an industry perspective. Neuropsychopharmacology 50, 258–268 (2025). URL https://www.nature.com/articles/s41386-024-01970-8.

[11] Law, E. & Chatfield, K. The undue burdens of clinical trial participation: implications for equity, diversity, and inclusion. Trials 27, 115 (2026). URL https://pmc.ncbi.nlm.nih.gov/articles/PMC12882127/.

[12] Meehan, A. J. et al. Clinical prediction models in psychiatry: a systematic review of two decades of progress and challenges. Molecular Psychiatry 27, 2700–2708 (2022). URL https://www.nature.com/articles/s41380-022-01528-4.

[13] Poldrack, R. A., Huckins, G. & Varoquaux, G. Establishment of Best Practices for Evidence for Prediction: A Review. JAMA Psychiatry 77, 534–540 (2020). URL 10.1001/jamapsychiatry.2019.3671.

[14] Vapnik, V. & Vashist, A. A new learning paradigm: Learning using privileged information. Neural Networks 22, 544–557 (2009). URL https://www.sciencedirect.com/science/article/pii/S0893608009001130.

[15] Vapnik, V. & Izmailov, R. Learning Using Privileged Information: Similarity Control and Knowledge Transfer. Journal of Machine Learning Research 16, 2023–2049 (2015). URL http://jmlr.org/papers/v16/vapnik15b.html.

[16] Vapnik, V. & Izmailov, R. Knowledge transfer in SVM and neural networks. Annals of Mathematics and Artificial Intelligence 81, 3–19 (2017). URL 10.1007/s10472-017-9538-x.

[17] Hinton, G., Vinyals, O. & Dean, J. Distilling the Knowledge in a Neural Network (2015). URL http://arxiv.org/abs/1503.02531. ArXiv:1503.02531 [stat.ML].

[18] Lopez-Paz, D., Bottou, L., Schölkopf, B. & Vapnik, V. Unifying distillation and privileged information (2016). URL http://arxiv.org/abs/1511.03643. ArXiv:1511.03643 [stat.ML].

[19] Martínez-García, M., Inza, I. & Lozano, J. A. Teacher privileged distillation: How to deal with imperfect teachers? Knowledge-Based Systems 316, 113338 (2025). URL https://www.sciencedirect.com/science/article/pii/S0950705125003855.

[20] Provodin, D., Akker, B. v. d., Katsimerou, C., Kaptein, M. & Pechenizkiy, M. Rethinking Knowledge Transfer in Learning Using Privileged Information (2024). URL http://arxiv.org/abs/2408.14319. ArXiv:2408.14319 [cs.LG].

[21] Stanton, S., Izmailov, P., Kirichenko, P., Alemi, A. A. & Wilson, A. G. Does Knowledge Distillation Really Work?, Vol. 34, 6906–6919 (Curran Associates, Inc., 2021). URL https://proceedings.neurips.cc/paper_files/paper/2021/hash/376c6b9ff3bedbbea56751a84fffc10c-Abstract.html.

[22] Franke, K., Ziegler, G., Klöppel, S., Gaser, C. & Alzheimer’s Disease Neuroimaging Initiative. Estimating the age of healthy subjects from T1-weighted MRI scans using kernel methods: exploring the influence of various parameters. NeuroImage 50, 883–892 (2010).

[23] Cole, J. H. & Franke, K. Predicting Age Using Neuroimaging: Innovative Brain Ageing Biomarkers. Trends in Neurosciences 40, 681–690 (2017).

[24] Cole, J. H. et al. Brain age predicts mortality. Molecular Psychiatry 23, 1385–1392 (2018). URL https://www.nature.com/articles/mp201762.

[25] Cole, J. H., Marioni, R. E., Harris, S. E. & Deary, I. J. Brain age and other bodily ‘ages’: implications for neuropsychiatry. Molecular Psychiatry 24, 266–281 (2019). URL https://www.nature.com/articles/s41380-018-0098-1.

[26] de Lange, A. G. et al. Mind the gap: Performance metric evaluation in brain-age prediction. Human Brain Mapping 43, 3113–3129 (2022). URL https://pmc.ncbi.nlm.nih.gov/articles/PMC9188975/.

[27] Gao, X. et al. Accelerated biological aging and risk of depression and anxiety: evidence from 424,299 UK Biobank participants. Nature Communications 14, 2277 (2023). URL https://www.nature.com/articles/s41467-023-38013-7.

[28] Qiu, W., Chen, H., Kaeberlein, M. & Lee, S.-I. ExplaiNAble BioLogical Age (ENABL Age): an artificial intelligence framework for interpretable biological age. The Lancet Healthy Longevity 4, e711–e723 (2023). URL https://www.thelancet.com/journals/lanhl/article/PIIS2666-7568%2823%2900189-7/fulltext.

[29] Bycroft, C. et al. The UK Biobank resource with deep phenotyping and genomic data. Nature 562, 203–209 (2018). URL https://www.nature.com/articles/s41586-018-0579-z.

[30] Schulz, M.-A., Siegel, N. T. & Ritter, K. Brain-age models with lower age prediction accuracy have higher sensitivity for disease detection. PLOS Biology 23, e3003451 (2025). URL https://journals.plos.org/plosbiology/article?id=10.1371/journal.pbio.3003451.

[31] Smith, S. M., Vidaurre, D., Alfaro-Almagro, F., Nichols, T. E. & Miller, K. L. Estimation of brain age delta from brain imaging. NeuroImage 200, 528–539 (2019).

[32] de Lange, A.-M. G. & Cole, J. H. Commentary: Correction procedures in brain-age prediction. NeuroImage : Clinical 26, 102229 (2020). URL https://pmc.ncbi.nlm.nih.gov/articles/PMC7049655/.

[33] Park, W., Kim, D., Lu, Y. & Cho, M. Relational Knowledge Distillation (2019). URL http://arxiv.org/abs/1904.05068. ArXiv:1904.05068 [cs.CV].

[34] Azad, R., Dehghanmanshadi, M., Khosravi, N., Cohen-Adad, J. & Merhof, D. Addressing Missing Modality Challenges in MRI Images: A Comprehensive Review. Computational Visual Media 11, 241–268 (2025). URL https://ieeexplore.ieee.org/document/10984423.

[35] Zhu, S. et al. Gee, J. C. et al. (eds) Bridging the Gap in Missing Modalities: Leveraging Knowledge Distillation and Style Matching for Brain Tumor Segmentation. (eds Gee, J. C. et al.) Medical Image Computing and Computer Assisted Intervention – MICCAI 2025, 95–106 (Springer Nature Switzerland, Cham, 2026).

[36] Kwak, M. G. et al. A Cross-Modal Mutual Knowledge Distillation Framework for Alzheimer’s Disease Diagnosis: Addressing Incomplete Modalities. IEEE Transactions on Automation Science and Engineering 22, 14218–14233 (2025). URL https://ieeexplore.ieee.org/document/10945821.

[37] Wang, M. et al. Robust multi-modal fusion architecture for medical data with knowledge distillation. Computer Methods and Programs in Biomedicine 260, 108568 (2025).

[38] Xing, X., Chen, Z., Hou, Y. & Yuan, Y. Gradient modulated contrastive distillation of low-rank multi-modal knowledge for disease diagnosis. Medical Image Analysis 88, 102874 (2023).

[39] Argelaguet, R. et al. MOFA+: a statistical framework for comprehensive integration of multi-modal single-cell data. Genome Biology 21, 111 (2020). URL 10.1186/s13059-020-02015-1.

[40] Zhao, Z., Pendiyala, K., Mortazavi, M. & Yan, N. PULSE: Privileged Knowledge Transfer from Rich to Deployable Sensors for Embodied Multi-Sensory Learning (2026). URL http://arxiv.org/abs/2510.24058. ArXiv:2510.24058 [eess.SP].

[41] Schöll, M. et al. PET Imaging of Tau Deposition in the Aging Human Brain. Neuron 89, 971–982 (2016).

[42] Benjamini, Y. & Hochberg, Y. Controlling the False Discovery Rate: A Practical and Powerful Approach to Multiple Testing. Journal of the Royal Statistical Society Series B: Statistical Methodology 57, 289–300 (1995). URL https://academic.oup.com/jrsssb/article/57/1/289/7035855.

[43] Kroenke, K., Spitzer, R. L. & Williams, J. B. W. The PHQ-9. Journal of General Internal Medicine 16, 606–613 (2001). URL 10.1046/j.1525-1497.2001.016009606.x.

[44] Spitzer, R. L., Kroenke, K., Williams, J. B. W. & Löwe, B. A brief measure for assessing generalized anxiety disorder: the GAD-7. Archives of Internal Medicine 166, 1092–1097 (2006).

[45] Bush, K., Kivlahan, D. R., McDonell, M. B., Fihn, S. D. & Bradley, K. A. The AUDIT alcohol consumption questions (AUDIT-C): an effective brief screening test for problem drinking. Ambulatory Care Quality Improvement Project (ACQUIP). Alcohol Use Disorders Identification Test. Archives of Internal Medicine 158, 1789–1795 (1998).

[46] Wiegert, J. et al. Breaking the norm: population-scale deviations of brain structure in depression and anxiety. Molecular Psychiatry 1–12 (2026). URL https://www.nature.com/articles/s41380-026-03691-4.

[47] Miller, K. L. et al. Multimodal population brain imaging in the UK Biobank prospective epidemiological study. Nature Neuroscience 19, 1523–1536 (2016). URL https://www.nature.com/articles/nn.4393.

[48] Alfaro-Almagro, F. et al. Image processing and Quality Control for the first 10,000 brain imaging datasets from UK Biobank. NeuroImage 166, 400–424 (2018).

[49] Fischl B. FreeSurfer. NeuroImage 62, 774–781 (2012).

[50] Desikan, R. S. et al. An automated labeling system for subdividing the human cerebral cortex on MRI scans into gyral based regions of interest. NeuroImage 31, 968–980 (2006).

[51] Schaefer, A. et al. Local-Global Parcellation of the Human Cerebral Cortex from Intrinsic Functional Connectivity MRI. Cerebral Cortex 28, 3095–3114 (2018).

[52] Yeo, B. T. T. et al. The organization of the human cerebral cortex estimated by intrinsic functional connectivity. Journal of Neurophysiology 106, 1125–1165 (2011).

[53] Townsend, P., Phillimore, P. & Beattie, A. Health and Deprivation: Inequality and the North (Routledge, London, 2023).

[54] Manard, M., Carabin, D., Jaspar, M. & Collette, F. Age-related decline in cognitive control: the role of fluid intelligence and processing speed. BMC neuroscience 15, 7 (2014).

[55] Squarzoni, P. et al. Relationship between regional brain volumes and cognitive performance in the healthy aging: an MRI study using voxel-based morphometry. Journal of Alzheimer’s disease: JAD 31, 45–58 (2012).

[56] Jiang, R. et al. Associations between grip strength, brain structure, and mental health in *>* 40,000 participants from the UK Biobank. BMC medicine 20, 286 (2022).

[57] Hamer, M. & Batty, G. D. Association of body mass index and waist-to-hip ratio with brain structure: UK Biobank study. Neurology 92, e594–e600 (2019).

[58] Stolicyn, A. et al. Comprehensive assessment of sleep duration, insomnia, and brain structure within the UK Biobank cohort. Sleep 47, zsad274 (2024).

[59] Lotze, M. et al. Income is associated with hippocampal/amygdala and education with cingulate cortex grey matter volume. Scientific Reports 10, 18786 (2020). URL https://www.nature.com/articles/s41598-020-75809-9.

[60] Cole, J. H. Multimodality neuroimaging brain-age in UK biobank: relationship to biomedical, lifestyle, and cognitive factors. Neurobiology of Aging 92, 34–42 (2020).

[61] Promet, L. et al. Brain Age Disparities in Psychosis Across DSM Diagnoses and B-SNIP Biotypes. Schizophrenia Bulletin 52, sbaf022 (2026).

[62] Mitchell, O., et al. Depression and aging: insights from brain age prediction models. Psychological Medicine 56, e204 (2026). URL https://www.cambridge.org/core/journals/psychological-medicine/article/depression-and-aging-insights-from-brain-age-prediction-models/8FE0690F7D1AEB3414ED28564FBECD00.

[63] Jawinski, P. et al. Genome-wide analysis of brain age identifies 59 associated loci and unveils relationships with mental and physical health. Nature Aging 5, 2086–2103 (2025). URL https://www.nature.com/articles/s43587-025-00962-7.

[64] Steiger, J. H. Tests for comparing elements of a correlation matrix. Psychological Bulletin 87, 245–251 (1980).

