## Supplementary Material for "Privilege Is Not Enough: Evaluating Generalized Distillation for Normative Brain Age Modeling in Depression, Anxiety and Alcohol Use Disorder"

### S1 Supplementary Methods

#### S1.1 Details for the normative cohort simulation

##### S1.1.1 Generative model and notation

For each participant $i$ we drew a chronological age $c_i \sim \mathcal{U}(20, 80)$ in years and two additive brain-aging offsets: a generic, symptom-independent offset $u_i$ and a symptom-linked offset $v_i$. True brain age was $\mathrm{BA}_i = c_i + u_i + v_i$ and the latent brain-age gap was $u_i + v_i$. In healthy controls $v$ was centered on zero and independent of severity; in cases $v$ increased with symptom severity $s$, so that the gap scaled with $s$.

The eight standard features were a noisy linear mixture of two latent channels - an age channel $p_i = c_i + u_i$ and a symptom channel $q_i = v_i$,

$$x_{ij} = W_j^{(1)}\, p_i + W_j^{(2)}\, q_i + \varepsilon_{ij}, \tag{S1}$$

with fixed loadings $W^{(1)}$ for the age channel and $W^{(2)}$ for the symptom channel and independent Gaussian noise $\varepsilon_{ij}$. The single privileged feature measured true brain age directly, $x_i^* = \mathrm{BA}_i + \varepsilon_i^*$. The symptom channel therefore is present in $x$ but only through the small loadings $W^{(2)}$, whereas $x^*$ carries it undiluted. Numeric values for all quantities are listed in Supplementary Section S1.1.3.

##### S1.1.2 Rationale for the two-channel construction

The construction isolates the mechanism of interest. The symptom-linked offset $v$ is embedded in the standard features $x$ through the loadings $W^{(2)}$ (Eq. S1), so it is in principle recoverable from $x$; however, because $v$ is uncorrelated with chronological age, the least-squares fit of the age target places no weight on the $v$-carrying feature directions, which instead behave as noise to be averaged away. A baseline model trained on chronological age is therefore effectively blind to the symptom signal, and its brain-age gap reflects only the recoverable age-channel offset. The teacher's soft targets equal its recovered brain age ($\approx c + u + v$) and explicitly reward the student for reading the weak symptom-channel directions that $x$ does contain.

This is why the transfer is possible at all; GD cannot inject information absent from the student's inputs, but it can teach the student to exploit a weak, otherwise-ignored signal that its features already carry. Were $v$ entirely absent from $x$ (privileged in the strict sense), no distillation scheme could recover it.

##### S1.1.3 Parameter settings

The generative model used $c \sim \mathcal{U}(20, 80)$ and $u \sim \mathcal{N}(0, 4^2)$ for all participants. The symptom-linked offset was $v \sim \mathcal{N}(0, 4^2)$ in controls and $v = 1 + 0.8\,s + \mathcal{N}(0, 1.5^2)$ in cases, with severity $s \sim \mathcal{U}(0, 10)$; true brain age was $\mathrm{BA} = c + u + v$. For the standard features (dimension $d = 8$), the age-channel and symptom-channel loadings were $W_j^{(1)} \sim \mathcal{U}(0.7, 1.3)$ and $W_j^{(2)} \sim \mathcal{U}(-1, 1) \times 1.3$, drawn once from a fixed seed and reused for every cohort, with measurement noise $\varepsilon_{ij} \sim \mathcal{N}(0, 3^2)$. The privileged feature used $\varepsilon_i^* \sim \mathcal{N}(0, 1^2)$. Each of the 50 repetitions drew 300 training controls, 500 held-out controls and 500 cases. The symptom channel's small variance relative to the age channel, whose spread is set by the 20–80-year age range, places the symptom signal in the weak-but-present regime that the rationale above requires; its loadings ($|W^{(2)}| \leq 1.3$) are comparable in size to those of the age channel.

##### S1.1.4 Model architectures and optimization

Throughout the simulation runs, the teacher was a scikit-learn MLP [1] regressor with one hidden layer of 32 ReLU units, trained on the standardized privileged feature trained with Adam [2] for up to 1,000 iterations, with all remaining settings at their scikit-learn defaults. The baseline and student were identical single-hidden-layer perceptrons (32 ReLU units, He initialization) implemented in PyTorch and trained by full-batch Adam (learning rate $10^{-2}$) for 400 epochs on standardized inputs and target. Feature and target standardization used training-control means and s.d. only, and the BAG was computed after de-standardizing the predicted age. No early stopping, regularization search, architecture search or learning-rate search was performed; the identical configuration was used for every condition and repetition, so that differences are attributable to the training signal ($x^*$ vs $x$; chronological-age labels vs distilled targets) rather than to tuning. The teacher's soft targets for the student were generated OOF, i.e. the training controls were split into five folds and each fold's soft brain ages were predicted by a teacher fitted on the remaining folds. To compare the student's predictions to those of the best-case model's, i.e. the teacher's, the non-deployable model was retained.

#### S1.2 Generalized distillation

Generalized distillation [3] extends the KD framework of [4] to the LUPI setting by using privileged features as the teacher's input modality. The procedure has three stages. In the analyses with UK Biobank data, age is treated throughout as an ordinal target over a shared grid of $K$ age bins $\{a_1, \ldots, a_K\}$ spaced at one year and spanning the training ages. A chronological age $y_i$ is encoded as a Gaussian-smoothed probability vector $t_i^{\text{age}}$ over the grid,

$$t_{i,k}^{\text{age}} \propto \exp\left(-\frac{(a_k - y_i)^2}{2\sigma^2}\right), \qquad \sum_k t_{i,k}^{\text{age}} = 1, \tag{S2}$$

with width $\sigma = 1$ year, and any model's predicted age is read out as the expectation $\hat{y} = \sum_k a_k \,\text{softmax}(z)_k$ of its logits $z$ over the grid.

The teacher, student and baseline are multilayer perceptrons (MLPs) that map their input features to logits over the age grid, each an MLP encoder followed by a linear age-bin head with layer normalization and dropout. Because the input dimensionality differs substantially across the three modalities (structural MRI, resting-state fMRI and the questionnaire battery), the teacher and student were not constrained to a shared architecture. Their depth, width and dropout, together with the optimizer learning rate and weight decay, were configured per modality and selected on the healthy-control development data, so each scenario uses its own teacher and student (Table S1). All models were trained with the Adam optimizer (decoupled weight decay) and gradient-norm clipping, with the number of epochs fixed to the cross-validated median of the validation-selected optimum. The baseline is architecturally and procedurally identical to the student and differs only in using $\lambda = 0$, so it never accesses the teacher. Any difference between student and baseline therefore reflects the distillation term alone and not an architectural difference.

##### S1.2.1 Soft label distillation

***Stage 1: Teacher training.***

A teacher model $f_t$ is trained on the privileged training data $\{(x_i^*, y_i)\}_{i=1}^{n_{\text{train}}}$ to map $x^*$ to logits over the age grid. It is fit by minimizing the cross-entropy between its softmax output and the Gaussian age target $t_i^{\text{age}}$ of Eq. S2. Both $x^*$ and $x$ are standardized independently before use.

***Stage 2: Soft label generation.***

For each training example the teacher produces logits over the age grid, which are converted into *soft labels* by a temperature $T > 1$ (here $T = 2$) that flattens the distribution,

$$q_i^{(T)} = \text{softmax}\big(z_i^t / T\big)\,, \tag{S3}$$

where $z_i^t = f_t(x_i^*)$ are the teacher's grid logits. Higher temperatures spread probability mass onto neighboring age bins, exposing the teacher's relative uncertainty about which ages are plausible for each subject.

***Stage 3: Student training.***

A student MLP is trained on the standard features $x$ using a weighted combination of two losses,

$$\mathcal{L} = (1-\lambda)\ \underbrace{H\big(t_i^{\text{age}},\ \text{softmax}(z_i)\big)}_{\text{age-supervision loss} \ \equiv\ \mathcal{L}_{\text{age}}}\ + \lambda\, T^2\, \underbrace{H\big(q_i^{(T)},\ \text{softmax}(z_i/T)\big)}_{\text{soft-label (distillation) loss}}, \tag{S4}$$

where $z_i$ are the student's logits over the age grid, $H(p, q) = -\sum_k p_k \log q_k$ is the cross-entropy, and $\lambda \in [0, 1]$ controls the relative weight of teacher imitation. The age-supervision term pulls the student toward the Gaussian age target and the distillation term pulls it toward the teacher's softened distribution at the same temperature. The $T^2$ factor compensates for the $1/T^2$ shrinkage of the soft-label gradient magnitude that arises when the logits are divided by $T$ [4]. Because both terms are cross-entropies over the same age grid, they are already on a comparable scale. Students were trained in minibatches of 64.

##### S1.2.2 Relational knowledge distillation

RKD is applied within the same three-stage procedure and to the same models described above; as before, the baseline uses $\lambda = 0$. The student objective differs from Eq. S4 only in the distillation term and in the absence of a temperature.

***Relational knowledge distillation loss.***

Soft-label distillation matches the teacher's predicted distribution for each subject independently. Relational knowledge distillation (RKD) [5] instead matches the geometry of the teacher's predictions across subjects. The two objectives share Stage 1 and the age-supervision term $\mathcal{L}_{\text{age}}$ of Eq. S4. Writing the brain-age gap of subject $i$ as $b_i = \hat{y}_i - y_i$, and given a minibatch of $B$ subjects, the relational term compares the within-batch pairwise BAG distances of student and teacher. For each pair $(i, j)$,

$$\psi^s_{ij} = \frac{|b^s_i - b^s_j|}{\mu_s}, \qquad \psi^t_{ij} = \frac{|b^t_i - b^t_j|}{\mu_t}, \tag{S5}$$

where $\mu_s$ and $\mu_t$ are the mean pairwise distances over the batch, a normalization that makes the term invariant to the overall scale of each model's BAGs. The distillation loss is the Huber (smooth-$L_1$, $\delta = 1$) discrepancy between the normalized distances, averaged over the $\binom{B}{2}$ pairs,

$$\mathcal{L}_{\text{RKD}} = \frac{1}{\binom{B}{2}} \sum_{i<j} h_\delta\left(\psi^s_{ij} - \psi^t_{ij}\right), \tag{S6}$$

with the teacher branch detached so that no gradient flows into it. The student objective replaces the soft-label term of Eq. S4 with $\mathcal{L}_{\text{RKD}}$,

$$\mathcal{L} = (1 - \lambda)\,\mathcal{L}_{\text{age}} + \lambda\, s_{\text{RKD}}\,\mathcal{L}_{\text{RKD}}, \tag{S7}$$

where $s_{\text{RKD}} = 50$ is a fixed multiplier that brings $\mathcal{L}_{\text{RKD}}$, which is of order $10^{-2}$, onto the scale of the supervised cross-entropy. It plays the role that the $T^2$ factor plays for soft labels in Eq. S4.

Two differences from soft-label distillation follow. First, relational distillation has no temperature parameter. The teacher enters only through the pairwise distances of its BAGs, so there is no output distribution to soften, and both the temperature $T$ and its $T^2$ gradient correction are absent, their scaling role taken by the fixed constant $s_{\text{RKD}}$. Second, only the relative structure of the teacher's BAGs is transferred and not their absolute values, because the distances are normalized by their batch mean. Matching the geometry of the BAGs rather than of predicted age concentrates the transfer on the teacher's age-deviation structure, which is the quantity of interest for the downstream associations.

***Batch size.***

Both the soft-label and relational students were trained with a batch size of 64. For soft-label distillation the loss is computed per subject and is insensitive to this choice. For relational distillation the target is instead defined over the $\binom{B}{2}$ pairs within a minibatch and each distance set is normalized by its within-batch mean, so a batch of 64 supplies $\binom{64}{2} = 2016$ pairwise relations per update and a stable normalizer. At least two subjects per batch are required for any pair to exist, and batches were drawn with `drop_last` enabled so that no update ran on a singleton final batch.

**Table S1**: **Model configurations for the relational-KD runs.** Each model is a MLP encoder followed by a linear projection to the representation dimension and a linear age-bin output head, with GELU activations, layer normalization and dropout. Architecture and optimization hyperparameters were selected per modality on the HC development folds; because the input dimensionality differs across sMRI, resting-state fMRI and the questionnaire battery, each scenario uses its own teacher and student. The baseline shares the student's architecture and optimization settings and differs only in the distillation weight ($\lambda = 0$); the student used $\lambda = 0.5$ for the BAG–outcome associations (Fig. 4) and $\lambda = 0.8$ for the transfer analyses (Fig. 5**b**,**c**). All runs used a batch size of 64, a Gaussian soft-label width $\sigma = 1$ year for teacher and student targets, one-year age bins, and five inner folds; the number of training epochs was fixed to the cross-validated median of the validation-selected optimum.

| **Scenario** | **Model** | **Encoder hidden** | **Rep. dim** | **Learning rate** | **Weight decay** | **Dropout** |
|---|---|---|---|---|---|---|
| Scenario 1 | Teacher | 128, 64 | 64 | $3 \times 10^{-4}$ | $1 \times 10^{-3}$ | 0.4 |
| (sMRI → questionnaire) | Student / Baseline | 16 | 8 | $3 \times 10^{-3}$ | $1 \times 10^{-3}$ | 0.4 |
| Scenario 2 | Teacher | 32 | 16 | $1 \times 10^{-3}$ | $1 \times 10^{-4}$ | 0.4 |
| (fMRI → sMRI) | Student / Baseline | 128, 64 | 64 | $3 \times 10^{-4}$ | $1 \times 10^{-3}$ | 0.4 |
| Scenario 3 | Teacher | 128, 64 | 64 | $3 \times 10^{-4}$ | $1 \times 10^{-3}$ | 0.4 |
| (sMRI → fMRI) | Student / Baseline | 32 | 16 | $3 \times 10^{-4}$ | $1 \times 10^{-3}$ | 0.4 |

#### S1.3 Imaging preparation and data filtering in UKB

Phenotype definitions and general data filtering followed Wiegert et al. [6]. Owing to the addition of rs-fMRI features (for scenarios 2 and 3) and a different optimization objective, subsequent sample sizes and correction for confounders differ from their work.

##### S1.3.1 Image acquisition

All imaging was acquired on identical Siemens Skyra 3T scanners with a 32-channel head coil across the UK Biobank dedicated imaging centers, following the standardized protocol [7, 8]. T1-weighted images were acquired with a sagittal 3D MPRAGE sequence at $1mm$ isotropic resolution ($TR = 2000$ ms, $TE = 2.01$ ms, $TI = 880$ ms, flip angle 8°, in-plane acceleration factor 2; ∼5 min). Resting-state fMRI was acquired with a multiband gradient-echo EPI sequence (multiband factor 8, no in-plane acceleration, fat saturation) at 2.4mm isotropic resolution, $TR/TE = 735/39$ ms, flip angle 52°, 490 volumes (∼6 min), with participants fixating a crosshair.

##### S1.3.2 Imaging-derived phenotypes and data fields

Structural features were the FreeSurfer image-derived phenotypes released by UK Biobank, comprising subcortical volumes from the automated subcortical segmentation (ASEG) and per-hemisphere cortical thickness and volume for the Desikan–Killiany atlas [9, 10]. Resting-state fMRI was obtained as the participant-level parcel time series bulk data (UK Biobank field 31018, imaging instance 2) and summarized into network-level connectivity (below). UK Biobank data-field identifiers for all feature sets are listed in Table S2.

| **Feature set** | **Regions** | **Measure** | **UK Biobank IDP field IDs** |
|---|---|---|---|
| Subcortical/CC volumes (ASEG) | 32 | volume | 26523, 26524, 26526, 26531–26535, 26554–26565, 26585–26596 |
| Cortical (Desikan–Killiany) | 33/hem. | thickness | lh 26756–26788; rh 26857–26889 |
| | 33/hem. | volume | lh 26789–26821; rh 26890–26922 |
| Temporal pole (lh/rh, thickness+volume) | 2 | — | — |

**Table S2**: **FreeSurfer structural features and corresponding UK Biobank imaging-derived-phenotype field IDs (for cross-reference).**

##### S1.3.3 Resting-state network connectivity

Node time series were extracted for the 100-parcel Schaefer parcellation, with each parcel assigned to one of the seven Yeo intrinsic connectivity networks [11, 12]. After discarding the first five non-steady-state volumes and z-scoring each parcel time series, a parcel-by-parcel Pearson correlation matrix was computed per participant. Correlations were then averaged within the network structure to yield seven within-network values (mean correlation among parcels of the same network) and 21 across-network values (mean correlation between the parcels of each network pair), giving 28 functional connectivity features, excluding self-correlations.

##### S1.3.4 Exclusion of neurodegenerative diseases

The exclusion criteria applied in this study (see Table S3) are the exact same as in Wiegert et al. [6]. We excluded $N = 2{,}589$ individuals with any ICD-10 code from Table S3.

##### S1.3.5 ICD-10 codes for psychiatric case and exclusion criteria

The ICD-10 codes used to define the three diagnostic case groups (MDD, ANX, AUD) and to exclude participants carrying other psychiatric diagnoses from the HC group are listed in Table S4. For MDD, single-episode (F32) and recurrent (F33) depressive diagnoses were combined, as the recurrent-only subgroup was too small to analyze separately; for all categories, every sub-code of the listed three-character blocks was included.

| ICD-10 Code | Condition |
|---|---|
| G30 | Alzheimer's Disease |
| G20 | Parkinson's Disease |
| G10 | Huntington's Disease |
| G12.2 | Amyotrophic Lateral Sclerosis (ALS) |
| G35 | Multiple Sclerosis |
| I60–I64 | Stroke |
| G45.9 | Transient Ischemic Attack (TIA) |
| S06 | Traumatic Brain Injury (TBI) |
| M79.7 | Fibromyalgia |
| G25.0 | Essential Tremor |
| G24 | Dystonia |
| G61.0 | Guillain-Barré Syndrome |
| G36.0 | Neuromyelitis Optica |
| B20 | HIV-associated neurocognitive disorders |
| C71 | Brain tumor |
| G91 | Hydrocephalus |

**Table S3**: **ICD-10 codes used to exclude participants with neurodegenerative, neurological, or related conditions from the UK Biobank instance two MRI cohort.** Conditions include major neurodegenerative diseases (e.g., Alzheimer's disease, Parkinson's disease, Huntington's disease, amyotrophic lateral sclerosis [ALS], multiple sclerosis), cerebrovascular events (stroke, transient ischemic attack [TIA]), traumatic brain injury (TBI), and other neurological disorders listed.

| ICD-10 Code | Condition | Use |
|---|---|---|
| F32–F33 | Major depressive disorder (single episode / recurrent) | MDD cases |
| F40–F41 | Phobic and other anxiety disorders | ANX cases |
| F10 | Mental and behavioural disorders due to alcohol use | AUD cases |
| F20 | Schizophrenia | HC exclusion |
| F31 | Bipolar affective disorder | HC exclusion |
| F90 | Hyperkinetic disorders (ADHD) | HC exclusion |
| F60.3 | Emotionally unstable (borderline) personality disorder | HC exclusion |

**Table S4**: **ICD-10 codes used to define psychiatric case groups and to exclude psychiatric diagnoses from the HC group in the UKB instance-two MRI cohort.** Case groups: major depressive disorder (MDD), anxiety disorder (ANX) and alcohol use disorder (AUD), each defined from all sub-codes of the listed blocks. HC exclusions are additional to the questionnaire-based cut-offs (PHQ-9 $< 5$, GAD-7 $< 5$, AUDIT-C $< 8$) described in Methods Section 3. Alzheimer's disease (G30) is additionally covered by the neurodegenerative-exclusion criteria in Table S3.

#### S1.4 Definition of alcohol consumption

In UK Biobank, alcohol units were defined as follows: pint or can of beer/lager/cider = 2 units; single shot of spirits (25 ml) = 1 unit; small glass of fortified wine = 1 unit; standard glass of wine (175 ml) = 2 units; large glass of wine (250 ml) = 3 units; bottle of wine (75 cl) = 9 units [6].

#### S1.5 UKB questionnaire fields

| Field ID | Showcase Description | Missing Value Indicators |
|---|---|---|
| p120112 | PHQ-9 item | ”Prefer not to answer” or null |
| p120111 | PHQ-9 item | ”Prefer not to answer” or null |
| p120110 | PHQ-9 item | ”Prefer not to answer” or null |
| p120109 | PHQ-9 item | ”Prefer not to answer” or null |
| p120108 | PHQ-9 item | ”Prefer not to answer” or null |
| p120107 | PHQ-9 item | ”Prefer not to answer” or null |
| p120106 | PHQ-9 item | ”Prefer not to answer” or null |
| p120105 | PHQ-9 item | ”Prefer not to answer” or null |
| p120104 | PHQ-9 item | ”Prefer not to answer” or null |

**Table S5**: **UK Biobank field IDs corresponding to items of the PHQ-9 (Patient Health Questionnaire-9) depression scale.** Each field represents a single questionnaire item. Missing values are coded as either ”Prefer not to answer” or left as null entries in the dataset.

| Field ID | Showcase Description | Missing Value Indicators |
|---|---|---|
| p29058 | GAD-7 item | ”Prefer not to answer” or null |
| p29059 | GAD-7 item | ”Prefer not to answer” or null |
| p29060 | GAD-7 item | ”Prefer not to answer” or null |
| p29061 | GAD-7 item | ”Prefer not to answer” or null |
| p29062 | GAD-7 item | ”Prefer not to answer” or null |
| p29063 | GAD-7 item | ”Prefer not to answer” or null |
| p29064 | GAD-7 item | ”Prefer not to answer” or null |

**Table S6**: **UK Biobank field IDs corresponding to items of the GAD-7 (Generalized Anxiety Disorder-7) scale.** Each field represents a single questionnaire item. Missing values are coded as either ”Prefer not to answer” or left as null entries in the dataset.

| Field ID | Description | Missing Value Indicators |
|---|---|---|
| p20414 | Alcohol consumption frequency | ”Prefer not to answer” or null |
| p20403 | Number of drinks consumed on a drinking day | ”Prefer not to answer” or null |
| p29093 | Frequency of consuming six or more units of alcohol | ”Prefer not to answer” or null |

**Table S7**: **UK Biobank field IDs corresponding to items of the AUDIT-C (Alcohol Use Disorders Identification Test-Consumption).** Each field represents one questionnaire item. Missing values are coded as either ”Prefer not to answer” or left as null entries.

##### S1.5.1 Rationale and construction details for the biosocial dataset

The variables selected to construct the scenario 1 questionnaire feature set were chosen according to four criteria. First, each is an established correlate of chronological age: performance in cognitive domains such as fluid intelligence and processing/reaction speed declines steadily across adulthood [13], and grip strength, adiposity, sleep and cardiometabolic measures likewise change systematically with age. Second, all of the selected measures have documented associations with brain structure—cognitive performance with regional gray-matter volume [14], grip strength with grey- and white-matter metrics [15], body mass index and waist-to-hip ratio with global and regional brain volume [16], sleep duration and insomnia with gray-matter volume [17], and socioeconomic indicators such as education and income with limbic and cortical gray-matter volume [18]; taken together, such biomedical, lifestyle and cognitive factors have been shown to relate to neuroimaging-derived brain age [19]. Third, all fields are recorded for a substantial fraction of participants, ensuring adequate overlap with the imaging sub-sample. Finally, every measure is obtained from questionnaire responses or simple physical measurements, and can therefore be collected at low cost and with minimal participant burden, without imaging, laboratory assays or specialist assessment.

We derived the scenario 1 questionnaire feature set used for the student and baseline model from the fields listed in Table S8. Because most phenotypes are recorded at more than one assessment visit, each predictor was resolved by an instance-priority fallback: for a given participant the value was taken from the imaging visit (instance 2) where available, and otherwise from the first repeat visit (instance 1) or, as a last resort, the baseline visit (instance 0).

The three cognitive measures – fluid intelligence, reaction time and pairs matching (instance 2) – were carried forward as continuous scores. The prospective memory field was binarized so that a correct recall on either the first or second attempt was collapsed into a single ”correct” category, distinguishing successful recall from failure to recall the instruction.

Lifestyle and health variables–smoking status, sleep duration, insomnia, body mass index and doctor-diagnosed diabetes – were retained directly after missing-code cleaning. A single waist-to-hip ratio was computed as the quotient of waist and hip circumference, with both measurements required to originate from the same assessment instance (again following the $2 \to 1 \to 0$ priority); non-finite values arising from the division were set to missing.

Hand grip strength was reduced to a single dominant-hand measure derived from the left and right dynamometer readings in conjunction with self-reported handedness. For right-handed participants the right-hand value was used, for left-handed participants the left-hand value, and for participants reporting use of both hands the larger of the two readings. Where handedness was missing or otherwise unspecified, the maximum of the left and right readings was taken. The grip readings and the handedness field were required to come from the same instance, applying the same instance-priority order.

Two ordinal sociodemographic variables were re-expressed on more interpretable scales. Educational qualifications were reverse-coded so that increasing values correspond to increasing attainment, and the categorical household income band was mapped to the approximate mid-point of each band in pounds sterling (£12,000, £24,500, £41,500, £76,000 and £125,000 for bands 1–5, respectively). The Townsend deprivation index [20] and number in household were retained after cleaning without further transformation.

Finally, the per-chunk processed records were concatenated, any remaining infinite values were converted to missing, and – because complete cases were required for the downstream analysis – rows with a missing value in any retained field were removed by listwise deletion. The resulting table therefore contains only participants with fully observed values across all questionnaire and physiological features.

| Field ID | Description | Missing Value Indicators |
|---|---|---|
| 20016-2.0 | Fluid intelligence score | null |
| 20023-2.0 | Reaction time (mean time to identify matches) | null |
| 399-2.0 | Pairs matching (number of incorrect matches) | null |
| 20018-2.0 | Prospective memory result | $-1$, or null |
| 20116-2.0 | Smoking status | $-3$, or null |
| 46 | Hand grip strength (left) | null |
| 47 | Hand grip strength (right) | null |
| 1707 | Handedness | $-3$, or null |
| 1160-2.0 | Sleep duration | $-1$, $-3$, or null |
| 1200-2.0 | Sleeplessness / insomnia | $-3$, or null |
| 21001-2.0 | Body mass index (BMI) | null |
| 48-2.0 | Waist circumference | null |
| 49-2.0 | Hip circumference | null |
| 2443-2.0 | Diabetes diagnosed by doctor | $-1$, $-3$, or null |
| 6138-2.0 | Educational qualifications | $-3$, $-7$, or null |
| 22189-0.0 | Townsend deprivation index | null |
| 738-2.0 | Average total household income before tax | $-1$, $-3$, or null |
| 709-2.0 | Number in household | $-1$, $-3$, or null |

**Table S8**: **UK Biobank field IDs used to construct the questionnaire and physiological feature set.** The field-ID suffix denotes the assessment instance and array index (e.g. -2.0 = imaging visit, instance 2). During processing the UK Biobank special missing codes $-1$ ("Do not know"), $-3$ ("Prefer not to answer") and $-7$ ("None of the above") were uniformly recoded as missing; remaining gaps are stored as null entries.

#### S1.6 Deconfounding

##### S1.6.1 Effect of deconfounding on the imaging features

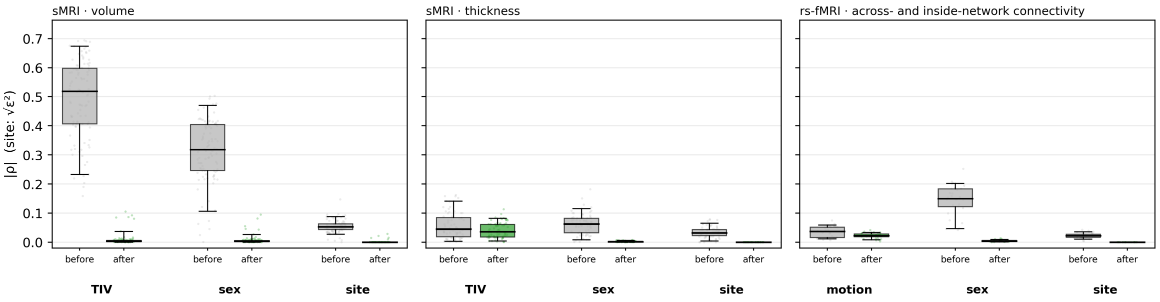


**Fig. S1**: **Feature-confounder associations before and after deconfounding.** Distributions of absolute Spearman rank correlation coefficients between the imaging data and confounders before and after deconfounding. Confounders include sex, site and total intracranial volume (TIV) in the case of volumetric sMRI freesurfer features and mean absolute head motion in the case of across- and inside-network rs-fMRI features.

##### S1.6.2 Definition of association model and transfer statistics

The semi-partial correlation residualized only the bias-corrected BAG on the covariate design $[\text{age}, \text{age}^2, \text{sex}, \text{age} \times \text{sex}, \text{age}^2 \times \text{sex}, \text{site}_{1,\cdots,k}, \text{TIV}, \text{mean motion}]$ (age mean-centered within each frame, site entered as the full indicator block, motion correction is used only in the Structural-functional sample) and correlated the residual with the untransformed outcome; intervals used the Fisher-$z$ transform with $n-k-3$ degrees of freedom and were cross-checked against HC3 heteroscedasticity-robust regressions of the outcome on the BAG. For the $\lambda$ sweep, the student was trained at $\lambda_1 = 0.5$ and $\lambda_2 = 0.8$ on identical subjects, and the two dependent correlations $r_{S,0.5}$ and $r_{S,0.8}$ of outcome $S$ with each student BAG were compared by the overlapping form of the Steiger test [21], given their mutual correlation $r_{\lambda_1,\lambda_2}$. $Z$ is computed as $z_{0.8} - z_{0.5}$, so a positive $Z$ indicates a stronger association at $\lambda = 0.8$. Recovery fractions and their bootstrap intervals resampled subjects jointly across the three models so the coefficients moved together on each draw, were re-residualized per resample, and were computed only for outcomes with a positive teacher advantage; where that advantage was small, as for the cognitive measures, the ratio was numerically unstable and the intervals are correspondingly wide. The cross-model comparisons of Fig. S4 used one-sided paired bootstraps (5000 resamples) on the raw BAGs. FDR-correction was applied within each family of tests, and every analysis used identical subject sets and standardization across teacher, baseline and student.

#### S1.7 Geometric visualization of the cross-model BAG correlations

Fig. 3 expresses the pairwise raw-BAG correlations described in Section 3 as a distance geometry, following the correlative-distance representation of the correlations between three variables [22]. Centering each model's raw BAGs on the group's shared complete-case frame and scaling them to unit Euclidean norm places each of teacher (T), student (S) and baseline (B) as a point on the unit sphere in an $n$-dimensional space, also termed person space [23], which has one axis per subject. In this space the Pearson correlation of two models is the cosine of the angle between their points [23], and their Euclidean distance is

$$d(i,j) = \sqrt{2\left(1 - r_{ij}\right)}. \tag{S8}$$

Eq. S8 is the correlative distance $\sqrt{2n(1-r_{ij})}$ of $z$-scored variables [22] divided by $\sqrt{n}$, which removes its dependence on sample size and makes edge lengths comparable across groups of different size. A shorter edge therefore corresponds to a higher correlation. Because all three correlations are computed on the same subjects, the three distances satisfy the triangle inequality, and because any three points in Euclidean space lie in a common plane, the triangle embeds in two dimensions with all pairwise distances preserved. We place the teacher at the origin, the student on the positive $x$-axis at distance $d(T,S)$ and the baseline by trilateration from the remaining two distances, and center each triangle on its centroid. All panels share identical axis limits and an equal aspect ratio, so a unit of length is the same in every subplot and a larger triangle denotes weaker overall agreement among the three models. Each edge is labeled with its Pearson $r$, the quantity on which all tests operate, and the student-teacher edge is highlighted in red. Because $d$ is nonlinear in $r$, a given difference in $r$ produces a larger change in edge length when correlations are high, so comparisons should rest on the edge labels.

Significance annotations reproduce the teacher-likeness test of Section 3. For each group and scenario a one-sided paired bootstrap (5,000 resamples, the same resampled subjects entering both correlations so their covariance is respected) tested whether the student is more teacher-like than the baseline (formally $r_{\mathrm{ST}} > r_{\mathrm{BT}}$). The resulting $p$-values were corrected across the family of all group × scenario cells by the Benjamini-Hochberg procedure, and asterisks on the student-teacher edge mark the corrected significance ($^{*}\, p_{FDR} < 0.05$, $^{**}\, p_{FDR} < 0.01$, $^{***}\, p_{FDR} < 0.001$). The panels are arranged with the HCs and the three ICD-10 diagnostic subgroups (MDD, ANX, AUD) as rows and the three scenarios as columns.

#### S1.8 Signal recovery and localization of the transferred component

Within each diagnostic subgroup let $n$ be the sample size and let $\mathbf{C} \in \mathbb{R}^{n\times(k+1)}$ be the confounder design matrix used throughout (intercept, mean-centered age and age$^2$, sex and their interactions, site indicators, TIV, mean absolute head motion (corrected only in the Structural-functional sample); $k$ denotes the number of non-constant columns). Write $\mathbf{R} = \mathbf{I} - \mathbf{C}(\mathbf{C}^\top\mathbf{C})^{+}\mathbf{C}^\top$ for the residual-maker that projects onto the orthogonal complement of the confounder space, and define the confounder-residualized BAGs as

$$\mathbf{g}_{\text{base}} = \mathbf{R}\,\text{BAG}_{\text{baseline}}, \qquad \mathbf{g}_{\text{stud}} = \mathbf{R}\,\text{BAG}_{\text{student}}. \tag{S9}$$

Both vectors are mean-zero because $\mathbf{C}$ contains the intercept. Let $\mathbf{s}$ denote the (raw) symptom-severity score.

##### S1.8.1 The distillation added component

The distillation-added component is the part of the residualized student BAG that is orthogonal to the residualized baseline BAG. Regressing $\mathbf{g}_{\text{stud}}$ on $[\mathbf{1},\, \mathbf{g}_{\text{base}}]$ and keeping the residual,

$$\mathbf{a} = (\mathbf{I} - \mathbf{P}_b)\,\mathbf{g}_{\text{stud}}, \qquad \mathbf{P}_b = \text{projection onto } \operatorname{span}\{\mathbf{1}, \mathbf{g}_{\text{base}}\}, \qquad \mathbf{a} \perp \mathbf{g}_{\text{base}}. \tag{S10}$$

Equivalently $\mathbf{g}_{\text{stud}} = \beta\,\mathbf{g}_{\text{base}} + \mathbf{a}$ with $\beta = \langle \mathbf{g}_{\text{stud}}, \mathbf{g}_{\text{base}}\rangle / \|\mathbf{g}_{\text{base}}\|^2$.

##### S1.8.2 Reported correlations

Each $r$ is a semi-partial correlation (predictor residualized on the confounders)

$$r_{\mathrm{B}} = \operatorname{corr}(\mathbf{g}_{\text{base}}, \mathbf{s}), \quad r_{\mathrm{S}} = \operatorname{corr}(\mathbf{g}_{\text{stud}}, \mathbf{s}), \quad r_{\text{add}} = \operatorname{corr}(\mathbf{a}, \mathbf{s}). \tag{S11}$$

$r_{\text{add}}$ is precisely the semi-partial correlation of severity with the student BAG *controlling for the baseline BAG.*

##### S1.8.3 Decomposition identity

Because $\mathbf{a} \perp \mathbf{g}_{\text{base}}$ we have $\sigma_a = \sigma_{\text{stud}}\sqrt{1-\rho^2}$ with $\rho = \text{corr}(\mathbf{g}_{\text{base}}, \mathbf{g}_{\text{stud}})$, and $\text{cov}(\mathbf{g}_{\text{stud}}, \mathbf{s}) = \beta\,\text{cov}(\mathbf{g}_{\text{base}}, \mathbf{s}) + \text{cov}(\mathbf{a}, \mathbf{s})$. Dividing by $\sigma_{\text{stud}}\sigma_s$ gives the exact relation

$$r_{\text{stud}} = \rho\, r_{\text{base}} + \sqrt{1-\rho^2}\, r_{\text{add}}, \tag{S12}$$

i.e. the raw student correlation is a norm-preserving blend ($\rho^2 + (1-\rho^2) = 1$) of the baseline and added correlations.

##### S1.8.4 Why $r_{add}$ can equal or exceed $r_S$

Eq. (S12) shows there is no ordering $r_S \geq r_{\text{add}}$ in general; the reverse is the standard signature of a suppressor. Rearranging, $r_{\text{add}} \geq r_S$ iff $r_B \leq r_{\text{add}}\,(1 - \sqrt{1-\rho^2})/\rho$. Two regimes produce it: (i) *dilution*, i.e. when the baseline BAG carries little severity signal ($r_B \approx 0$), Eq. (S12) reduces to $r_S = \sqrt{1-\rho^2}\, r_{\text{add}} \leq r_{\text{add}}$, because the baseline-shared variance is a severity-irrelevant component in the denominator of $r_S$ but is removed by construction from $\mathbf{a}$; and (ii) *sign cancellation*, which occurs when $r_B$ opposes $r_{\text{add}}$, which drives $r_S$ strictly below $r_{\text{add}}$.

Combining both regimes clarifies the intended reading of Fig. 5**c**. Where transfer occurs, the severity signal is concentrated in the distillation-added component rather than inherited from the non-distilled baseline.

##### S1.8.5 Inference

$p$-values and 95% confidence intervals for $r_{\text{add}}$ use the Fisher z-transformation [24], $z = \text{artanh}(r_{\text{add}})$, with effective degrees of freedom $\nu = n - k - 4$, accounting for the $k$ confounder terms plus the intercept and the baseline component removed before correlating; the interval is $\tanh\left(z \pm z_{1-\alpha/2}/\sqrt{\nu}\right)$. This is mildly conservative.

##### S1.8.6 Contextualization of the observed values

Table S15 reports the six symptom-subgroup panels. In both highlighted positive-transfer panels (Scenario 1/GAD-7, Scenario 3/AUDIT-C) the baseline correlation is essentially zero, so by Eq. (S12) $r_{\text{add}} \geq r_S$ (the dilution regime). In every panel where the baseline already carries signal ($r_B$ = 0.06–0.14) the inequality reverses, exactly as Eq. (S12) requires. No panel exhibits sign cancellation ($r_B$ is non-negative throughout).

### S2 Supplementary Results

#### S2.1 Analyses on simulated data

Fig. S2 depicts the teacher-likeness of the student and the baseline measured via correlations of their raw (age-uncorrected) BAGs derived from simulated data. The student model's BAGs are uniformly more teacher-like than the baseline's, but the correlation between the student's and baseline's BAGs is comparable to the student-teacher correlation on HC's and even more elevated in simulated cases.

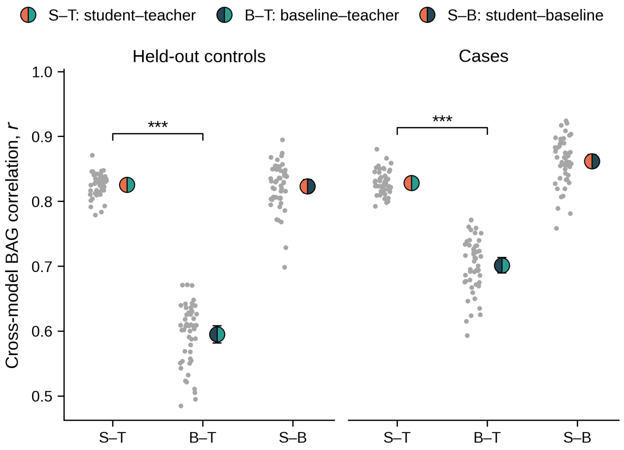


**Fig. S2**: **Cross-model BAG correlations in the simulation.** Pearson correlations of uncorrected BAGs between the teacher, student and baseline models across simulated HCs and cases. The weight between age supervision and teacher imitation ($\lambda = 0.8$) was the same as in the rest of the analyses with the simulated data. Brackets denote significant differences between correlation pairs (*** $p_{FDR} < 0.001$).

##### S2.1.1 BAG-severity coupling across models on simulated data

| **Model** | **r(BAG, sev.)** | **Slope (pt/y)** | $\mathbf{BAG_{HC}}$ | $\mathbf{BAG_{Case}}$ | **Case-HC sep.** |
|---|---|---|---|---|---|
| Teacher | $0.418 \pm 0.045$ | $+0.696 \pm 0.081$ | +0.01 | +4.32 | $+4.31 \pm 0.36$ |
| Baseline | $-0.008 \pm 0.062$ | $-0.011 \pm 0.092$ | +0.01 | −0.26 | $-0.27 \pm 0.45$ |
| Student | $0.217 \pm 0.043$ | $+0.356 \pm 0.077$ | +0.04 | +2.12 | $+2.08 \pm 0.35$ |

**Table S9**: **BAG-severity association on simulated data.** The privileged teacher shows a strong BAG–severity coupling and clear case elevation; the deployable baseline ($x$ only, chronological-age labels) recovers neither; the distilled student recovers a substantial fraction of both while, like the baseline, using only $x$ at deployment. Slope is severity-points per year of brain-age gap (BAG as predictor, severity as dependent variable).

##### S2.1.2 Summary of the three analysis steps on simulated data

| **Hypothesis (paired across splits)** | **Effect [95% CI]** | $d_z$ | $p_{\mathrm{FDR}}$ | **Sig.** |
|---|---|---|---|---|
| Student's teacher-likeness is greater than baseline's | $+0.127\,[+0.115, +0.139]$ (r) | 3.08 | $3.7 \times 10^{-27}$ | *** |
| Student's BAG-severity $r$ greater than baseline's | $+0.225\,[+0.212, +0.238]$ (r) | 4.93 | $2.2 \times 10^{-36}$ | *** |
| Student separates cases from controls (BAG sep $> 0$) | $+2.076\,[+1.975, +2.178]$ (yr) | 5.80 | $2.1 \times 10^{-39}$ | *** |

**Table S10**: **Results of hypothesis tests on simulated data.** Paired hypothesis tests across independent splits, one-sided $t$-tests with FDR-correction. $d_z$ is Cohen's $d$ for paired samples; effects are in Pearson correlation units (r) or years (yr). $^{***}p_{\mathrm{FDR}} < 0.001$.

##### S2.1.3 Robustness analyses

***Distillation weight***

Fig. S3 shows that sweeping $\lambda$ from 0 to 1 moved the student monotonically from the baseline toward the teacher on both the BAG-severity correlation (from $r \approx 0$ at $\lambda = 0$ to $r \approx 0.26$ at $\lambda = 1$) and the case-control BAG separation (from $\approx 0$ to $\approx 2.7$ years), establishing that the transfer is driven by the distillation term of the loss rather than by other elements of the pipeline. The headline analysis (Fig. 2) used $\lambda = 0.8$, at which the student still fits chronological age while imitating the teacher.

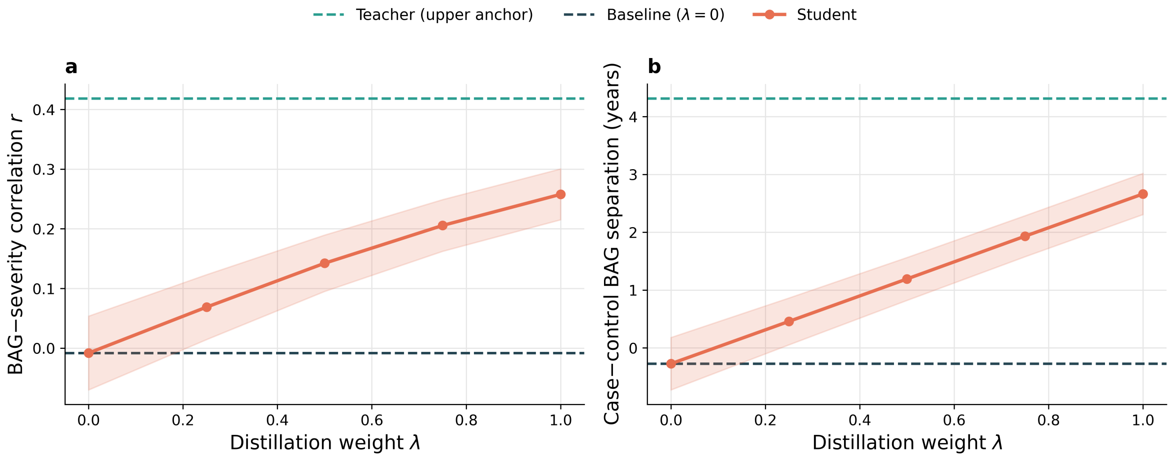


**Fig. S3**: **Transfer of the privileged brain-age gap as a function of the distillation weight $\lambda$. a,** the BAG-severity correlation and **b,** the case-control BAG separation. Points and line, mean over 50 repetitions; shaded band, ± s.d.; dashed lines, teacher and baseline references.

***Soft-target cross-fitting.***

Replacing the OOF teacher soft targets with in-sample teacher predictions left the student's BAG–severity correlation unchanged (student $r = 0.22$ in both cases; $r = 0.223$ versus 0.224 when the standard linear age-bias correction was additionally applied), confirming that the low-capacity, one-dimensional teacher does not overfit its training controls and that the reported transfer is not an artifact of in-sample soft labeling. This tolerance is consistent with recent evidence that soft labels need not match the ground-truth labels to train an effective student, and that even substantially biased teachers can yield good students [25].

###### S2.1.4 Scope and limitations

The teacher is the best-case model that retains privileged access at test time and is not deployable; the comparison of interest is student versus baseline, both of which use only $x$ at deployment. The synthetic symptom signal is linear and low-dimensional, and severity is independent of age by construction. The simulation is intended to establish the mechanism, i.e. that a privileged-information brain-age gap can be partially handed down to a standard-feature student through the distillation loss, and to motivate the empirical analysis, not to forecast an effect size on imaging data.

#### S2.2 Age-prediction performance metrics

| Scenario | Model (modality) | MAE (y) | RMSE (y) | $r$ | $n_{\text{test}}$ |
|---|---|---|---|---|---|
| 1 (sMRI→ qu.) | Teacher (sMRI) | 4.42 | 5.58 | 0.680 | 11,118 |
| | Student (questionnaire) | 6.31 | 7.60 | 0.113 | |
| | Baseline (questionnaire) | 5.45 | 6.73 | 0.488 | |
| 2 (fMRI → sMRI) | Teacher (rs-fMRI) | 5.98 | 7.26 | 0.278 | 17,477 |
| | Student (sMRI) | 6.03 | 7.25 | 0.519 | |
| | Baseline (sMRI) | 4.31 | 5.43 | 0.698 | |
| 3 (sMRI → fMRI) | Teacher (sMRI) | 4.30 | 5.44 | 0.699 | 17,477 |
| | Student (rs-fMRI) | 6.19 | 7.48 | 0.182 | |
| | Baseline (rs-fMRI) | 6.01 | 7.30 | 0.272 | |

**Table S11**: **Performance of the teacher, student and baseline models.** In each scenario, age prediction performance was computed on the raw predicted age (before bias correction) over all test data, i.e. the healthy-control normative holdout pooled with every case. MAE and RMSE are in years; $r$ is the Pearson correlation between raw predicted and chronological age. All three models within a scenario are trained on the exact same healthy control sample and scored on the identical complete-case test set of size $n_{\text{test}}$. Abbreviations: MAE, mean absolute error; RMSE, root mean squared error.

#### S2.3 Teacher-likeness analysis on UKB via cross-model BAG correlation

| Scenario | Group | $n$ | $r_{\text{ST}}$ | $r_{\text{BT}}$ | $r_{\text{SB}}$ | $p_{\text{ST>BT}}$ | | $p_{\text{ST>SB}}$ | |
|---|---|---|---|---|---|---|---|---|---|
| 1 | HC | 3566 | 0.681 | 0.593 | 0.880 | $< .001$ | *** | 1.000 | n.s. |
| | MDD | 400 | 0.716 | 0.645 | 0.895 | $< .001$ | *** | 1.000 | n.s. |
| | ANX | 371 | 0.667 | 0.613 | 0.904 | $< .001$ | *** | 1.000 | n.s. |
| | AUD | 595 | 0.682 | 0.628 | 0.893 | $< .001$ | *** | 1.000 | n.s. |
| 2 | HC | 6350 | 0.959 | 0.653 | 0.698 | $< .001$ | *** | $< .001$ | *** |
| | MDD | 715 | 0.963 | 0.707 | 0.735 | $< .001$ | *** | $< .001$ | *** |
| | ANX | 649 | 0.964 | 0.685 | 0.716 | $< .001$ | *** | $< .001$ | *** |
| | AUD | 935 | 0.960 | 0.655 | 0.696 | $< .001$ | *** | $< .001$ | *** |
| 3 | HC | 6350 | 0.633 | 0.620 | 0.978 | $< .001$ | *** | 1.000 | n.s. |
| | MDD | 715 | 0.679 | 0.676 | 0.980 | .289 | n.s. | 1.000 | n.s. |
| | ANX | 649 | 0.659 | 0.655 | 0.981 | .245 | n.s. | 1.000 | n.s. |
| | AUD | 935 | 0.638 | 0.626 | 0.979 | .019 | * | 1.000 | n.s. |

**Table S12**: **Cross-model raw-BAG correlations by scenario and group.** Underlying statistics for the geometric visualization (Figure 3) and the confidence-interval panels (Figure S4). $n$ is the sample size of each group; $r_{\text{ST}}$, $r_{\text{BT}}$ and $r_{\text{SB}}$ are the pairwise Pearson correlations of the raw brain-age gaps. $p_{\text{ST>BT}}$ and $p_{\text{ST>SB}}$ are Benjamini-Hochberg FDR-corrected $p$-values from one-sided paired bootstraps (5,000 resamples; the same resampled subjects entering both correlations), each corrected within its own family of twelve group × scenario cells. Groups are the ICD-10 diagnostic subgroups (MDD, ANX, AUD) and the HC normative holdout. Significance: $^{*}p_{FDR} < 0.05$, $^{**}p_{FDR} < 0.01$, $^{***}p_{FDR} < 0.001$.

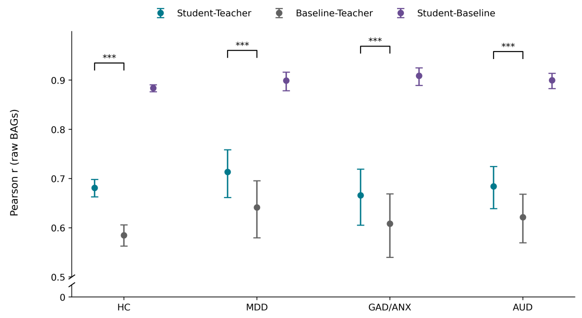


(a) Scenario 1

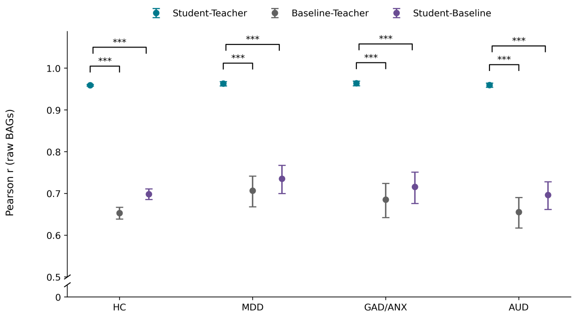


(b) Scenario 2

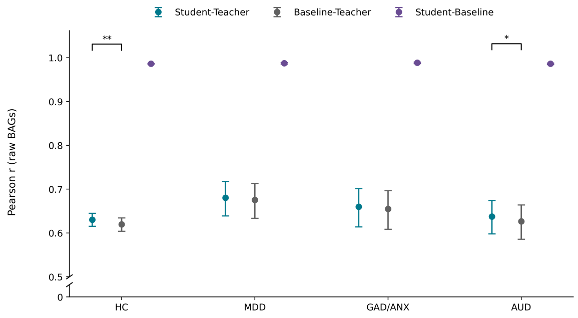


(c) Scenario 3

**Fig. S4**: **Cross-model raw-BAG correlations with confidence intervals and both directional tests** (extended version of Fig. 3). For each distillation scenario (panels) and group (HCs and the ICD-10 MDD, ANX and AUD diagnostic subgroups) the three pairwise Pearson correlations of the raw BAGs, student-teacher, baseline-teacher and student-baseline, are shown as points with 95% Fisher-$z$ confidence intervals on a broken $y$-axis. Significance brackets mark the two directional comparisons, $r_{\mathrm{ST}} > r_{\mathrm{BT}}$ (the student is more teacher-like than the baseline) and $r_{\mathrm{ST}} > r_{\mathrm{SB}}$, each assessed by a one-sided paired bootstrap (5,000 resamples, the same resampled subjects entering both correlations so their covariance is respected); brackets are drawn only where the comparison is significant after FDR correction ($^{*}\,p_{FDR} < 0.05$, $^{**}\,p_{FDR} < 0.01$, $^{***}\,p_{FDR} < 0.001$). This figure extends the visual information content of Figure 3 by additionally displaying the confidence intervals and the second directional test. Note that the Benjamini-Hochberg FDR correction here is applied within each scenario, across that scenario's family of group $\times$ directional-test comparisons, whereas the asterisks in the main-text Figure 3 correct the $r_{\mathrm{ST}} > r_{\mathrm{BT}}$ test across all twelve group $\times$ scenario cells; the two schemes can therefore assign slightly different significance levels to the same comparison.

#### S2.4 BAG-symptom severity association analysis for scenario 2

Fig. S5 shows the BAG-symptom severity associations of the teacher, student and baseline in scenario 2 at $\lambda = 0.5$, which we omitted from the main text as the rs-fMRI teacher showed no elevated association (beyond what was available from the deployable sMRI information) that the student could inherit.

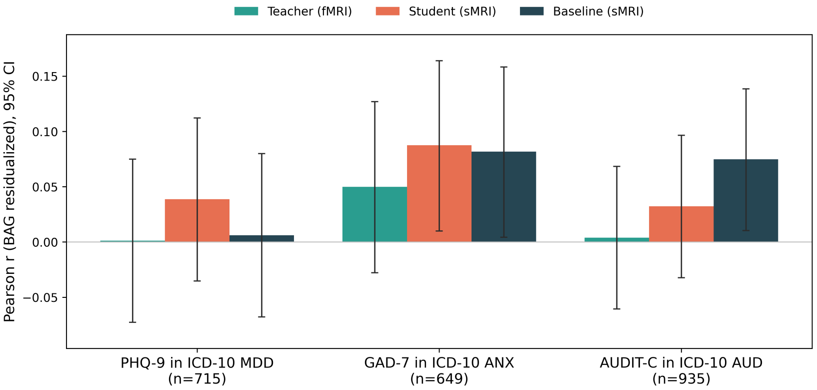


**Fig. S5**: **BAG–symptom-severity associations across models in scenario 2.** Scenario 2 (teacher rs-fMRI, student and baseline sMRI, $\lambda = 0.5$). BAG-symptom-severity correlations (residualized BAG, 95% CI) in the ICD-10 MDD, ANX and AUD subgroups.

#### S2.5 Effect of imitation weight increase on BAG-symptom severity association

Table S13 lists the effect of an increase in imitation strength on the student's BAG-outcome correlation in each UK Biobank scenario where we detected a transfer.

| **Comparison** | $n$ | $r_{\lambda_1}$ | $r_{\lambda_2}$ | $\Delta r$ | $r(\text{BAG}_{\lambda_1}, \text{BAG}_{\lambda_2})$ | $Z$ | $p$ | $p_{FDR}$ | **Sig.** |
|---|---|---|---|---|---|---|---|---|---|
| Scenario 1 – ANX | 371 | 0.053 | 0.097 | 0.044 | 0.932 | 2.27 | 0.011 | 0.034 | yes |
| Scenario 3 – AUD | 935 | 0.049 | 0.058 | 0.009 | 0.941 | 0.82 | 0.205 | 0.308 | no |
| Scenario 3 – RT | 7552 | 0.028 | 0.029 | 0.001 | 0.938 | 0.16 | 0.435 | 0.435 | no |

**Table S13**: **Results of the $\lambda$ sweep.** For each transferring association the residualized Pearson $r$ is compared between the balanced ($\lambda_1 = 0.5$) and transfer-heavy ($\lambda_2 = 0.8$) students on the same subjects. $r(\text{BAG}_{\lambda_1}, \text{BAG}_{\lambda_2})$ is the correlation between the two students' BAGs (the dependency term of the overlapping-correlations test). Significance is a one-sided Steiger test ($r_{\lambda=0.8} > r_{\lambda=0.5}$); raw and Benjamini-Hochberg corrected $p$-values across the rows. $Z$ is computed as $z_{0.8} - z_{0.5}$ (positive values indicate a stronger association at $\lambda = 0.8$). Abbreviations: ANX, anxiety disorder; AUD, alcohol use disorder; RT, reaction time.

#### S2.6 Signal recovery metrics

##### S2.6.1 Transfer of available teacher signal to student

**Table S14**: **Signal recovery across scenarios.** For each outcome the residualized Pearson $r$ of the teacher, baseline and student BAGs is shown, together with the teacher's advantage over baseline ($r_{\text{teacher}} - r_{\text{baseline}}$), the student's gain over baseline ($r_{\text{student}} - r_{\text{baseline}}$) and the recovery fraction (student gain divided by teacher advantage) with its bootstrap 95% CI. Symptom outcomes are evaluated in the matched diagnostic subgroups; cognitive outcomes in cases. Recovery fractions are unstable where the teacher advantage is small and their CIs are correspondingly wide. All student and baseline values are from the $\lambda = 0.8$ run.

| Scenario | Outcome | $n$ | $r_T$ | $r_B$ | $r_S$ | Teacher gap | Student gap | Recovery [95% CI] |
|---|---|---|---|---|---|---|---|---|
| **Scenario 1** | **GAD-7/ANX (symptom)** | 371 | 0.183 | 0.006 | **0.101** | 0.177 | 0.095 | 0.54 [−0.50, 1.70] |
| Scenario 1 | AUDIT-C/AUD (symptom) | 595 | 0.104 | 0.063 | 0.024 | 0.041 | −0.039 | −0.95 [−12.7, 12.6] |
| **Scenario 3** | **AUDIT-C/AUD (symptom)** | 935 | 0.080 | 0.003 | **0.055** | 0.078 | 0.052 | 0.67 [−0.90, 4.46] |
| Scenario 3 | GAD-7/ANX (symptom) | 649 | 0.088 | 0.045 | 0.037 | 0.043 | −0.008 | −0.19 [−4.16, 5.35] |
| Scenario 3 | Reaction time (cognition) | 7552 | 0.051 | 0.027 | 0.027 | 0.024 | 0.001 | 0.03 [−2.41, 1.74] |
| Scenario 3 | Pairs matching (cognition) | 7552 | 0.023 | 0.014 | 0.008 | 0.010 | −0.006 | −0.60 [−6.50, 6.94] |
| Scenario 3 | Fluid intelligence (cognition) | 7552 | −0.034 | −0.042 | −0.021 | 0.008 | 0.021 | 2.78 [−16.1, 15.5] |

##### S2.6.2 Distillation-added component

Table S15 decomposes each student BAG into the part shared with the non-distilled baseline and the orthogonal distillation-added component, and reports the semi-partial correlation of each with symptom severity. In the two subgroups showing positive transfer (bold), the severity association is carried almost entirely by the added component ($r_{add} \geq r_S$) while the baseline-shared part contributes little ($r_B \approx 0$); the reconstructed $\hat{r}_S = \rho\ r_B + \sqrt{1-\rho^2}\ r_{add}$ reproduces the observed $r_S$ to three decimals in every row, confirming the decomposition is exact.

**Table S15**: **Decomposition of the student brain-age gap in each matched diagnostic subgroup.** The confounder-residualized student BAG is split into the part shared with the non-distilled baseline BAG and the orthogonal *distillation-added* component; $r_{\mathrm{add}}$ is the semi-partial Pearson correlation of that added component with symptom severity (Fisher 95% CI and $p$, on $n-k-4$ degrees of freedom), and $r_B$, $r_S$ are the corresponding correlations of the baseline and full student BAGs. $\rho = \mathrm{corr}(g_{\mathrm{baseline}}, g_{\mathrm{student}})$ is the baseline–student coupling, and $\hat{r}_S = \rho\, r_B + \sqrt{1-\rho^2}\, r_{\mathrm{add}}$ reconstructs the student correlation from the decomposition. The two subgroups showing positive transfer are in bold. All student and baseline values are from the $\lambda = 0.8$ run. $p$ is uncorrected; BH correction across the two bold rows gives $p_{FDR} = 0.056$ (ANX) and 0.048 (AUD).

| Scenario | Outcome | $n$ | $k$ | $r_{\mathrm{add}}$ [95% CI] | $p$ | $r_B$ | $r_S$ | $\rho$ | $\hat{r}_S$ |
|---|---|---|---|---|---|---|---|---|---|
| Scenario 1 | PHQ-9 / MDD | 400 | 9 | 0.063 [−0.037, 0.161] | 0.217 | 0.136 | 0.071 | 0.064 | 0.071 |
| **Scenario 1** | **GAD-7 / ANX** | **371** | **9** | **0.101** [−0.003, **0.202**] | **0.056** | **0.006** | **0.101** | **0.122** | **0.101** |
| Scenario 1 | AUDIT-C / AUD | 595 | 9 | 0.017 [−0.065, 0.098] | 0.688 | 0.063 | 0.024 | 0.112 | 0.024 |
| Scenario 3 | PHQ-9 / MDD | 715 | 10 | 0.031 [−0.043, 0.105] | 0.412 | 0.023 | 0.038 | 0.678 | 0.038 |
| Scenario 3 | GAD-7 / ANX | 649 | 10 | 0.008 [−0.070, 0.085] | 0.844 | 0.045 | 0.037 | 0.695 | 0.037 |
| **Scenario 3** | **AUDIT-C / AUD** | **935** | **10** | **0.074 [ 0.010, 0.138]** | **0.024** | **0.003** | **0.055** | **0.707** | **0.055** |

### Supplementary References

[1] Rumelhart, D. E., Hinton, G. E. & Williams, R. J. Learning representations by back-propagating errors. *Nature* **323**, 533–536 (1986). URL https://www.nature.com/articles/323533a0.

[2] Kingma, D. P. & Ba, J. Adam: A Method for Stochastic Optimization. *CoRR* (2014). URL https://www.semanticscholar.org/paper/Adam%3A-A-Method-for-Stochastic-Optimization-Kingma-Ba/a6cb366736791bcccc5c8639de5a8f9636bf87e8.

[3] Lopez-Paz, D., Bottou, L., Schölkopf, B. & Vapnik, V. Unifying distillation and privileged information (2016). URL http://arxiv.org/abs/1511.03643. ArXiv:1511.03643 [stat.ML].

[4] Hinton, G., Vinyals, O. & Dean, J. Distilling the Knowledge in a Neural Network (2015). URL http://arxiv.org/abs/1503.02531. ArXiv:1503.02531 [stat.ML].

[5] Park, W., Kim, D., Lu, Y. & Cho, M. Relational Knowledge Distillation (2019). URL http://arxiv.org/abs/1904.05068. ArXiv:1904.05068 [cs.CV].

[6] Wiegert, J. *et al.* Breaking the norm: population-scale deviations of brain structure in depression and anxiety. *Molecular Psychiatry* 1–12 (2026). URL https://www.nature.com/articles/s41380-026-03691-4.

[7] Miller, K. L. *et al.* Multimodal population brain imaging in the UK Biobank prospective epidemiological study. *Nature Neuroscience* **19**, 1523–1536 (2016). URL https://www.nature.com/articles/nn.4393.

[8] Alfaro-Almagro, F. *et al.* Image processing and Quality Control for the first 10,000 brain imaging datasets from UK Biobank. *NeuroImage* **166**, 400–424 (2018).

[9] Fischl, B. FreeSurfer. *NeuroImage* **62**, 774–781 (2012).

[10] Desikan, R. S. *et al.* An automated labeling system for subdividing the human cerebral cortex on MRI scans into gyral based regions of interest. *NeuroImage* **31**, 968–980 (2006).

[11] Schaefer, A. *et al.* Local-Global Parcellation of the Human Cerebral Cortex from Intrinsic Functional Connectivity MRI. *Cerebral Cortex* **28**, 3095–3114 (2018).

[12] Yeo, B. T. T. *et al.* The organization of the human cerebral cortex estimated by intrinsic functional connectivity. *Journal of Neurophysiology* **106**, 1125–1165 (2011).

[13] Manard, M., Carabin, D., Jaspar, M. & Collette, F. Age-related decline in cognitive control: the role of fluid intelligence and processing speed. *BMC neuroscience* **15**, 7 (2014).

[14] Squarzoni, P. *et al.* Relationship between regional brain volumes and cognitive performance in the healthy aging: an MRI study using voxel-based morphometry. *Journal of Alzheimer's disease: JAD* **31**, 45–58 (2012).

[15] Jiang, R. *et al.* Associations between grip strength, brain structure, and mental health in $>$ 40,000 participants from the UK Biobank. *BMC medicine* **20**, 286 (2022).

[16] Hamer, M. & Batty, G. D. Association of body mass index and waist-to-hip ratio with brain structure: UK Biobank study. *Neurology* **92**, e594–e600 (2019).

[17] Stolicyn, A. *et al.* Comprehensive assessment of sleep duration, insomnia, and brain structure within the UK Biobank cohort. *Sleep* **47**, zsad274 (2024).

[18] Lotze, M. *et al.* Income is associated with hippocampal/amygdala and education with cingulate cortex grey matter volume. *Scientific Reports* **10**, 18786 (2020). URL https://www.nature.com/articles/s41598-020-75809-9.

[19] Cole, J. H. Multimodality neuroimaging brain-age in UK biobank: relationship to biomedical, lifestyle, and cognitive factors. *Neurobiology of Aging* **92**, 34–42 (2020).

[20] Townsend, P., Phillimore, P. & Beattie, A. *Health and Deprivation: Inequality and the North* (Routledge, London, 2023).

[21] Steiger, J. H. Tests for comparing elements of a correlation matrix. *Psychological Bulletin* **87**, 245–251 (1980).

[22] Vos, P. Pearson's correlation between three variables; using students' basic knowledge of geometry for an exercise in mathematical statistics. *International Journal of Mathematical Education in Science and Technology* **40**, 533–541 (2009). URL https://doi.org/10.1080/00207390802419578.

[23] Lee Rodgers, J. & Nicewander, W. A. Thirteen Ways to Look at the Correlation Coefficient. *The American Statistician* **42**, 59–66 (1988). URL https://doi.org/10.1080/00031305.1988.10475524.

[24] Fisher, R. A. Frequency Distribution of the Values of the Correlation Coefficient in Samples from an Indefinitely Large Population. *Biometrika* **10**, 507–521 (1915). URL https://www.jstor.org/stable/2331838.

[25] Yuan, H. *et al.* *Learning From Biased Soft Labels*, Vol. 36, 59566–59584 (Curran Associates, Inc., 2023). URL https://proceedings.neurips.cc/paper_files/paper/2023/hash/bad8ddaed5feb552f9e8f2e37c0531a1-Abstract-Conference.html.
